# Genomic Basis of Multiple Myeloma Subtypes from the MMRF CoMMpass Study

**DOI:** 10.1101/2021.08.02.21261211

**Authors:** Sheri Skerget, Daniel Penaherrera, Ajai Chari, Sundar Jagannath, David S. Siegel, Ravi Vij, Gregory Orloff, Andrzej Jakubowiak, Ruben Niesvizky, Darla Liles, Jesus Berdeja, Moshe Levy, Jeffrey Wolf, Saad Z. Usmani, The MMRF CoMMpass Network, Austin W. Christofferson, Sara Nasser, Jessica L. Aldrich, Christophe Legendre, Brooks Benard, Chase Miller, Bryce Turner, Ahmet Kurdoglu, Megan Washington, Venkata Yellapantula, Jonathan R. Adkins, Lori Cuyugan, Martin Boateng, Adrienne Helland, Shari Kyman, Jackie McDonald, Rebecca Reiman, Kristi Stephenson, Erica Tassone, Alex Blanski, Brianne Docter, Meghan Kirchhoff, Daniel C. Rohrer, Mattia D’Agostino, Manuela Gamella, Kimberly Collison, Jennifer Stumph, Pam Kidd, Andrea Donnelly, Barbara Zaugg, Maureen Toone, Kyle McBride, Mary DeRome, Jennifer Yesil, David Craig, Winnie Liang, Norma C. Gutierrez, Scott D. Jewell, John Carpten, Kenneth C. Anderson, Hearn Jay Cho, Daniel Auclair, Sagar Lonial, Jonathan J. Keats

**Affiliations:** Integrated Cancer Genomics Division, Translational Genomics Research Institute, Phoenix, AZ, 85004, USA; Tisch Cancer Institute, Icahn School of Medicine at Mount Sinai, New York, NY, 10029, USA; Hackensack University Medical Center, Hackensack, NJ, 07601, USA; Division of Oncology, Washington University, St. Louis, MO, 63110, USA; Virginia Cancer Specialists, Fairfax, VA, 22031, USA; University of Chicago Medical Center, Chicago, IL, 60637, USA; Weill Cornell Medicine, New York, NY, 10065, USA; Division of Hematology/Oncology, East Carolina University, Greenville, NC, 27834, USA; Sarah Cannon Research Institute, Nashville, TN, 37203, USA; Baylor Scott & White Research Institute, Dallas, TX, 75246, USA; Department of Medicine, UCSF Medical Center, San Francisco, CA, 94117, USA; Levine Cancer Institute, Charlotte, NC, 28204, USA; Multiple Myeloma Research Foundation, Norwalk, CT, 06851, USA; Van Andel Institute, Grand Rapids, MI, 49503, USA; Myeloma Unit, Division of Hematology, University of Torino, Azienda Ospedaliero-Universitaria Citt a della Salute e della Scienza di Torino, Torino, Italy; Spectrum Health, Grand Rapids, MI, 49503, USA; Precision for Medicine, Flemington, NJ, 08822, USA; InStat Services, Chatham, NJ, 07928, USA; Department of Hematology, University Hospital of Salamanca, IBSAL, CIBERONC, Salamanca, Spain; Dana-Farber Cancer Institute, Harvard Cancer Center, Boston, MA, 02215, USA; Department of Hematology and Medical Oncology, Emory University School of Medicine, Atlanta, GA, 30322, USA

## Abstract

Multiple myeloma is a treatable, but currently incurable, hematological malignancy of plasma cells characterized by diverse and complex tumor genetics for which precision medicine approaches to treatment are lacking. The MMRF CoMMpass study is a longitudinal, observational clinical study of newly diagnosed multiple myeloma patients where tumor samples are characterized using whole genome, exome, and RNA sequencing at diagnosis and progression, and clinical data is collected every three months. Analyses of the baseline cohort identified genes that are the target of recurrent gain- and loss-of-function events. Consensus clustering identified 8 and 12 unique copy number and expression subtypes of myeloma, respectively, identifying high-risk genetic subtypes and elucidating many of the molecular underpinnings of these unique biological groups. Analysis of serial samples showed 25.5% of patients transition to a high-risk expression subtype at progression. We observed robust expression of immunotherapy targets in this subtype, suggesting a potential therapeutic option.

## Introduction

Multiple myeloma is a treatable, but currently incurable, hematological malignancy of plasma cells. The incorporation of new treatment modalities over the last two decades vastly improved overall survival of myeloma patients, however, patients still ultimately relapse and there remains a subset of high-risk patients with poor outcomes. Despite significant efforts to understand the molecular basis of the disease, predicting patient outcomes and identifying high-risk patients remains a challenge.

Multiple myeloma is a genetically heterogeneous disease with two broad karyotypic groups. A hyperdiploid (HRD) phenotype, with characteristic trisomies of chromosomes 3, 5, 7, 9, 11, 15, 19, and 21, is present in 50-60% of tumors^1–3^. The remaining non-hyperdiploid (NHRD) tumors, with pseudo-diploid karyotypes, typically have an immunoglobulin translocation dysregulating NSD2/WHSC1/MMSET, MYC, CCND1, or MAF^4–9^. Tumors harbor many other genetic aberrations including non-immunoglobulin structural abnormalities and mutations^10–13^. Although previous genomic studies were instrumental in deconvoluting the genetic heterogeneity of myeloma, they are mostly limited by small cohort sizes, the number and types of assays performed, lack of longitudinal sampling, clinical follow up, and biased inclusion of heavily pretreated patients, limiting our comprehensive understanding of the disease.

To better understand the impact of tumor genetic profile on patient outcomes and treatment response, the Multiple Myeloma Research Foundation sponsored the Relating Clinical Outcomes in Multiple Myeloma to Personal Assessment of Genetic Profile (CoMMpass) Study (NCT01454297). CoMMpass is an ongoing, prospective, longitudinal, observational clinical study which accrued 1143 newly diagnosed, previously untreated multiple myeloma patients from sites throughout the United States, Canada, Spain, and Italy between 2011-2016. Comprehensive molecular profiling is performed on bone marrow derived tumor samples collected at diagnosis and each progression event using whole genome (WGS), whole exome (WES), and RNA (RNAseq) sequencing. Clinical parameters are collected every three months through the eight year observation period.

We present a molecular analysis of the complete baseline cohort, with a median follow up of 3 years, identifying recurrent loss- and gain-of-function events and distinct copy number (CN) and gene expression subtypes of myeloma. The comprehensive nature of this dataset and our integrated analysis framework defines both the overall frequency of gene alterations in myeloma and the genetic basis of a high-risk patient population that does not benefit from current therapies.

## Results

### Cohort Description

The CoMMpass cohort includes 1143 patients from 84 clinical sites located in the United States, Canada, Spain, and Italy. The demographic and clinical parameters of the cohort at diagnosis adhere to expected distributions (Tables 1 & S1). Median age at diagnosis was 63 (range 27-93) with an expected over representation of males (60.4%) versus females (39.6%). This cohort is largely composed of patients from the United States and, unlike most clinical trials, the distribution of self-reported ancestry reflects US Census Bureau statistics with 80.6% Caucasian, 17.5% Black, and 1.9% Asian. Baseline prognostication of patients with the international staging system (ISS) identified 35.1% ISSI, 35.1% ISSII, and 27.2% ISSIII^14^.

**Table 1.**
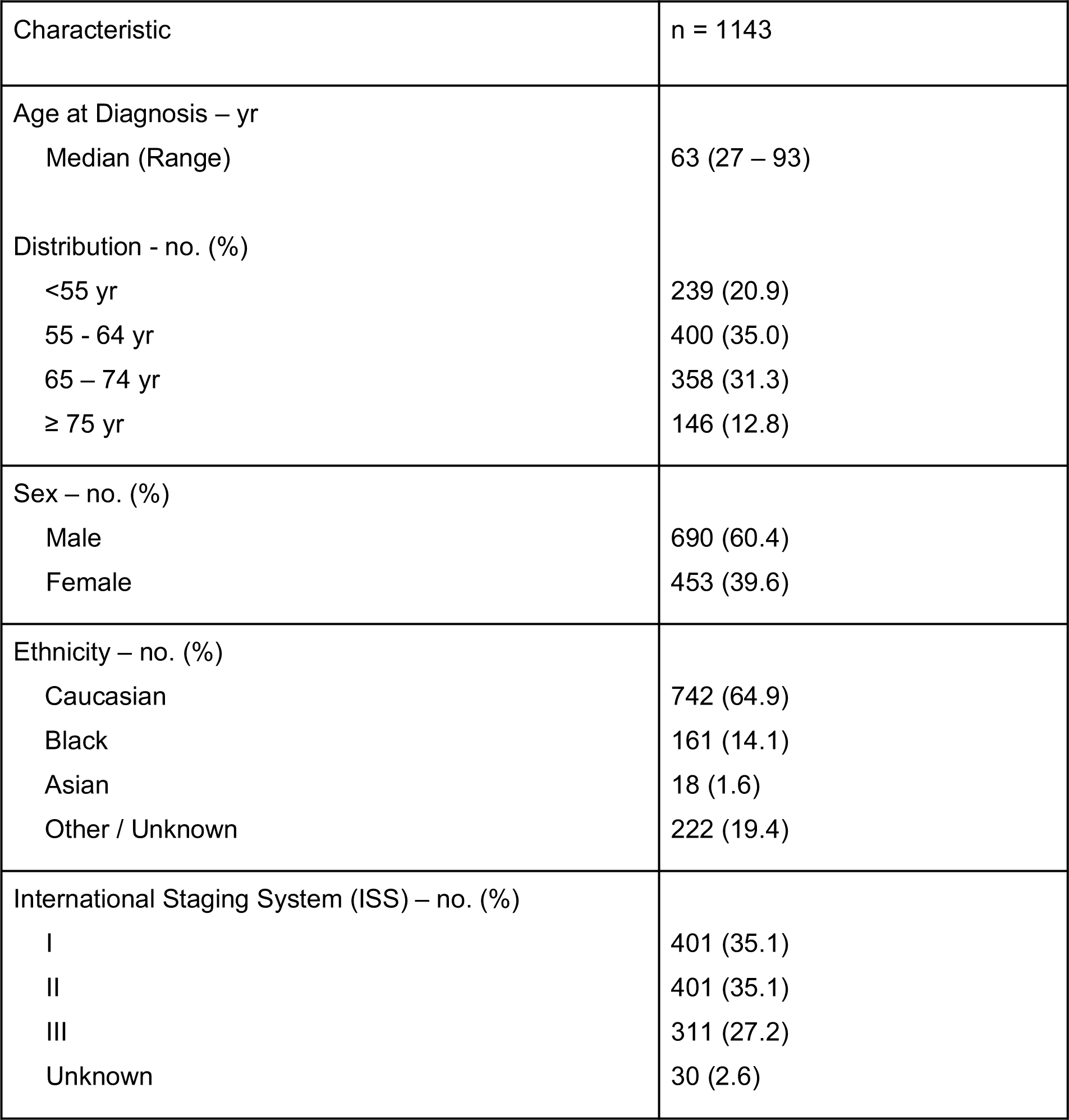
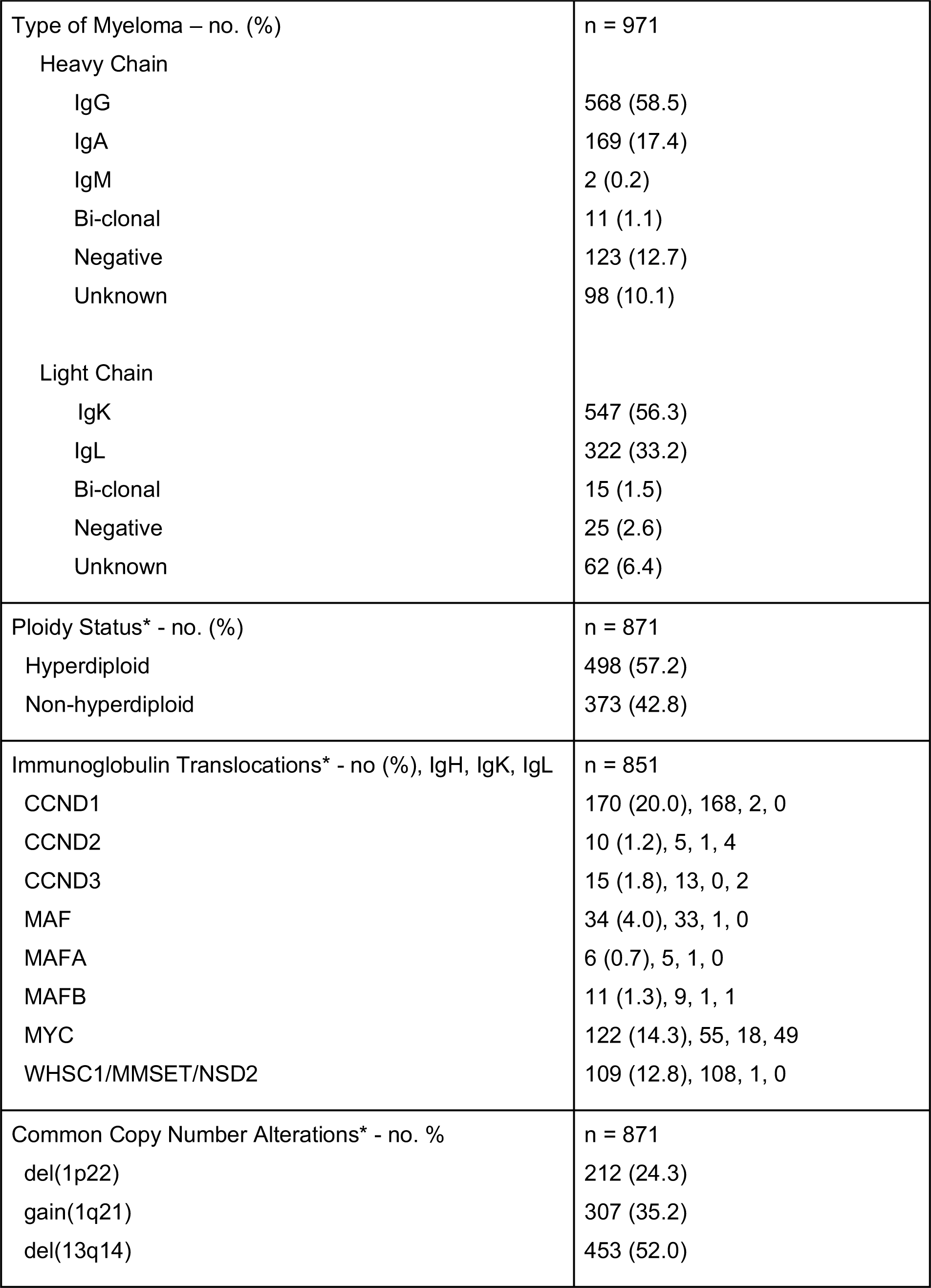
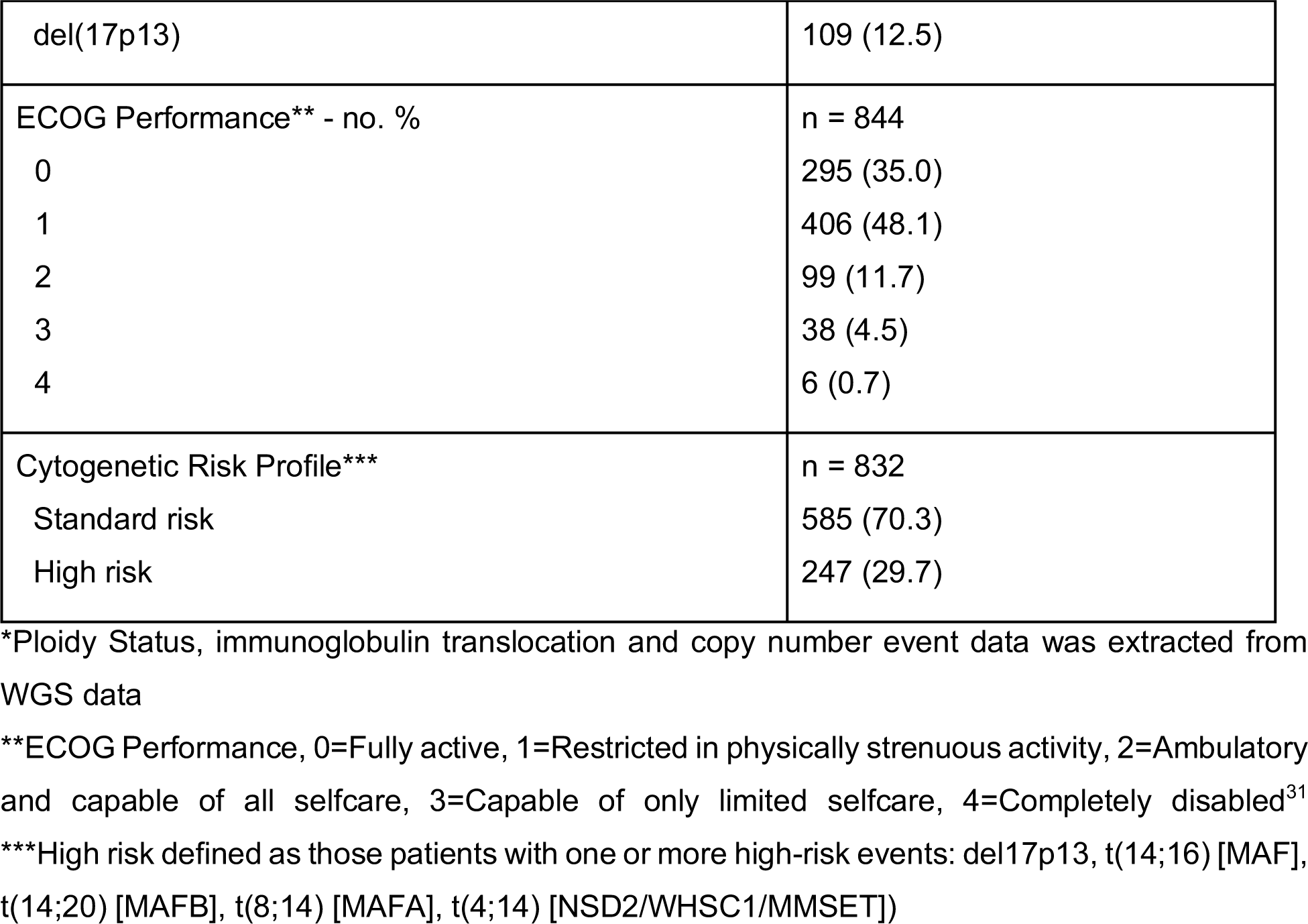
Characteristics of the baseline CoMMpass cohort

Due to the highly variable cytogenetic panels used at individual sites, we defined the phenotype of each patient from WGS data. This identified 57.2% HRD and 42.8% NHRD patients (based on the detection of a whole chromosome gain on at least two classic hyperdiploid chromosomes: 3, 5, 7, 9, 11, 15, 19, and 21), 24.3% patients with del(1p22), 35.2% with gain(1q21), 52.0% with del(13q14), and 12.5% with del(17p13). We identified translocations involving common target genes from any of the three immunoglobulin loci occurring at the following frequencies: 20.0% CCND1; 1.2% CCND2; 1.8% CCND3; 4.0% MAF; 0.7% MAFA; 1.3% MAFB; 14.3% MYC; and 12.8% WHSC1. Of these events, 83.0% involved the IgH locus while 5.3% and 11.7% involved the kappa and lambda IgL loci, respectively.

Irrespective of treatment, the median progression-free survival (PFS) of the cohort was 36 months and median overall survival (OS) of the cohort was 74 months (Figure S1.1A-B). Median OS for ISSIII patients was 54 months, while median OS for ISSI and ISSII patients could not be confidently predicted (Figure S1.1C-D). Patients with at least one high-risk cytogenetic feature had worse OS outcomes, even with uniform utilization of novel agents (Figure S1.2)^15^.

### Integrated Analysis for Gain and Loss of Function Genes

To comprehensively identify loss-of-function (LOF) and gain-of-function (GOF) events in myeloma patients, an integrated model was developed to overcome the limitations of analyzing any one data type by combining measurements from WES, WGS, and RNAseq to assign a functional state to each gene. In the LOF model, a single event in a gene was designated a partial LOF, whereas genes with two or more events were designated a complete LOF. At diagnosis, 592 patients had all three sequencing assays performed and were included in the analysis (Figure S1.3, Table S6). We identified at least one partial LOF in all patients, a complete LOF event in 92% of patients, and 70 genes where a complete LOF event was identified in ≥5 patients (Figure 1A). Complete LOF was observed in 12 genes in >2% of the cohort, including TRAF3 (10.1%), DIS3 (6.9%), FAM46C (5.1%), CYLD (4.7%), TP53 (4.1%), MAX (3.5%), RB1 (3.2%), WWOX (3.2%), HUWE1 (2.7%), PVT1 (2.5%), CDC42BPB (2.0%), and MAGEC1 (2.0%). However, CDC42BPB is unlikely a tumor suppressor gene in myeloma as it is in a contiguous gene region on chr14 with TRAF3, which was previously shown to be the target of bi-allelic loss in this region^16^.

**Figure 1.**
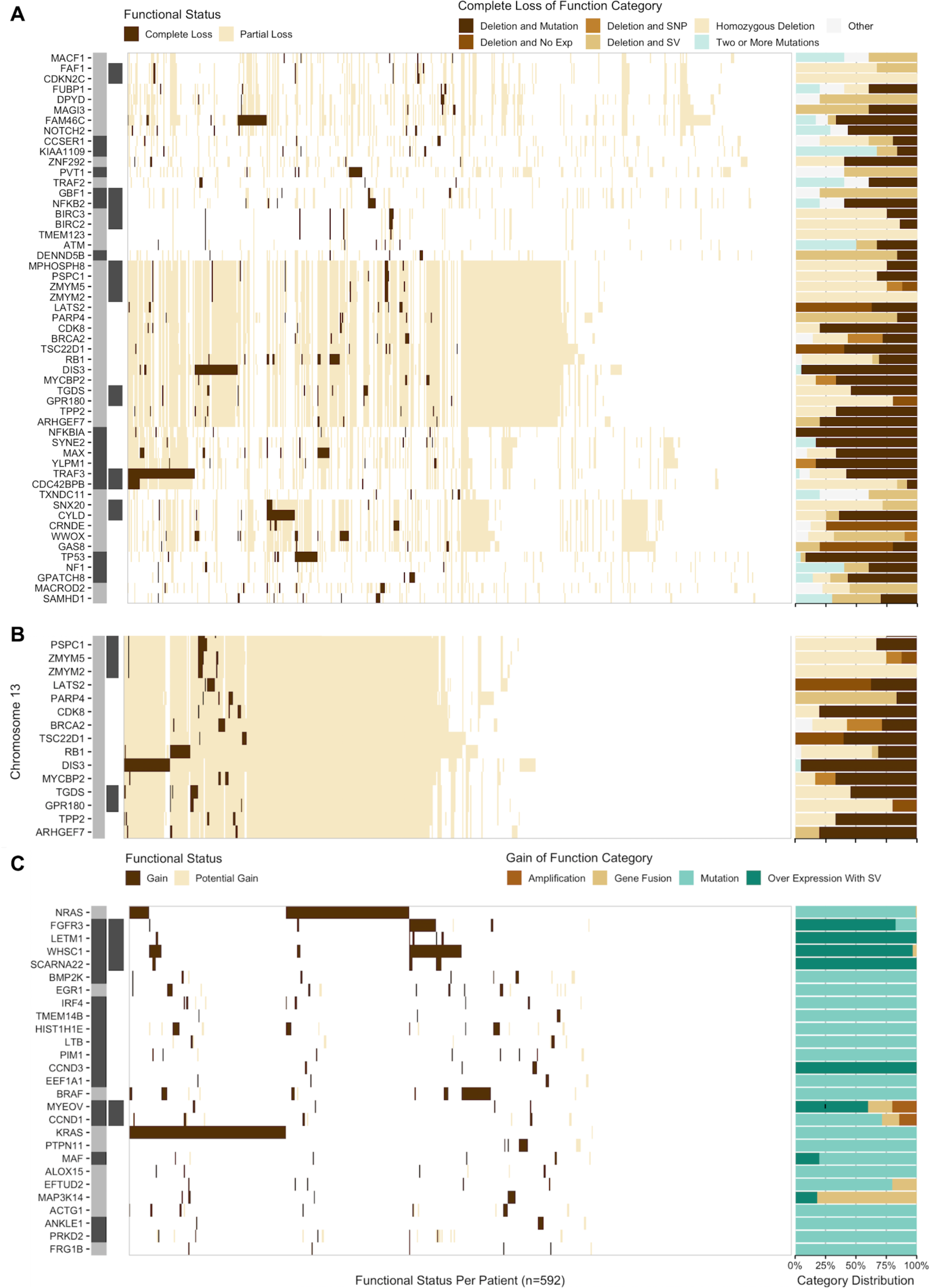
Recurrent LOF and GOF events occurring in at least five patients at diagnosis ordered by event frequency. The location and proximity of individual genes is shown next to each gene with the alternating gray and black bar illustrating when the chromosomal location changes, while black bars directly to the right denote contiguous genes. (A) Complete LOF was observed in 53 autosomal located genes. (B) Genes on chromosome 13q that were the target of complete LOF events in at least five patients in the baseline cohort. (C) GOF events were detected in 27 autosomal genes.

The target gene(s) of chr13 loss continues to be controversial. WGS data detected 13q14 deletion in 52.0% of patients, while LOF analysis identified 26.5% had complete LOF of one or more genes on chr13. The commonly assumed target, RB1, showed complete LOF in 3.2% while DIS3 complete LOF was detected in 6.9%, however, a striking number of additional genes were independently knocked-out in myeloma (Figure 1B). Two contiguous gene regions with complete LOF were identified: the first comprising MPHOSPH8 (1.4%), PSPC1 (1.5%), ZMYM5 (1.4%), ZMYM2 (1.0%); and the second comprising TGDS (1.9%) and GPR180 (0.8%) where the minimal region of deletion and LOF frequency suggest the targets are PSPC1 and TGDS, respectively. Additional complete LOF events were identified targeting LATS2 (1.4%), BRCA2 (1.2%), PARP4 (1.0%), MYCBP2 (1.0%), TPP2 (1%), CDK8 (0.8%), TSC22D1 (0.8%), and ARHGEF7 (0.8%).

These results highlight that monosomy 13 is associated with multiple independent gene inactivation events.

The GOF analysis identified an event in 92% of patients at diagnosis and 27 genes where a GOF event was identified in 5 or more patients (Figure 1C). There were 7 genes in which a GOF event was identified in greater than 2% of the cohort, including KRAS (23.6%), NRAS (21.6%), WHSC1 (10.3%), BRAF (7.1%), FGFR3 (4.9%), HIST1H1E (3.2%), and EGR1 (2.5%).

A number of patients had activating mutations described in other cancers including KRAS^G12D^ (3.2%), KRAS^G12V^ (1.5%), NRAS^Q61R^ (7.4%), NRAS^Q61K^ (5.9%), NRAS^Q13R^ (2.4%), NRAS^Q61H^ (2.0%), NRAS^Q13D^ (0.8%), and BRAF^V600E^ (3.9%).

### Identification of Copy Number Subtypes of Multiple Myeloma

To discover potential underlying phenotypes of myeloma beyond the known dichotomy of HRD and NHRD karyotypes, unsupervised consensus clustering was performed on CN data from 871 patients. Three independent replicates identified eight subtypes as the optimal solution (Figure S2.1). These trials were highly consistent, with deviations between any two replicates occurring in <1% of patients (Figures S2.2 & S2.3).

The CN subtypes consisted of five HRD and three NHRD subtypes and were annotated based on defining features (Figure 2A). The HRD, classic subtype had gains of classic HRD chromosomes, and the remaining HRD subtypes were annotated based on deviations from this phenotype. The HRD, ++15 subtype exhibited tetrasomy of chr15 (Figure S2.4) while two subtypes (HRD, diploid 7 and HRD, diploid 3, 7) were defined by the absence of chr7 and chr3 trisomies. Finally, the complex HRD, +1q, diploid 11, -13 subtype lacked chr11 trisomy but harbored gain of chr1q and loss of chr13. Of the NHRD subtypes, the diploid subtype was mostly devoid of CN events and highly associated with translocations targeting a D-type cyclin (71.3%). The remaining two NHRD groups were strongly associated with canonical immunoglobulin translocations (71.1%) and were defined by chr13 loss. The -13 subtype contained a subpopulation of patients with chr14 loss, while the +1q, -13 subtype had gains of 1q.

**Figure 2.**
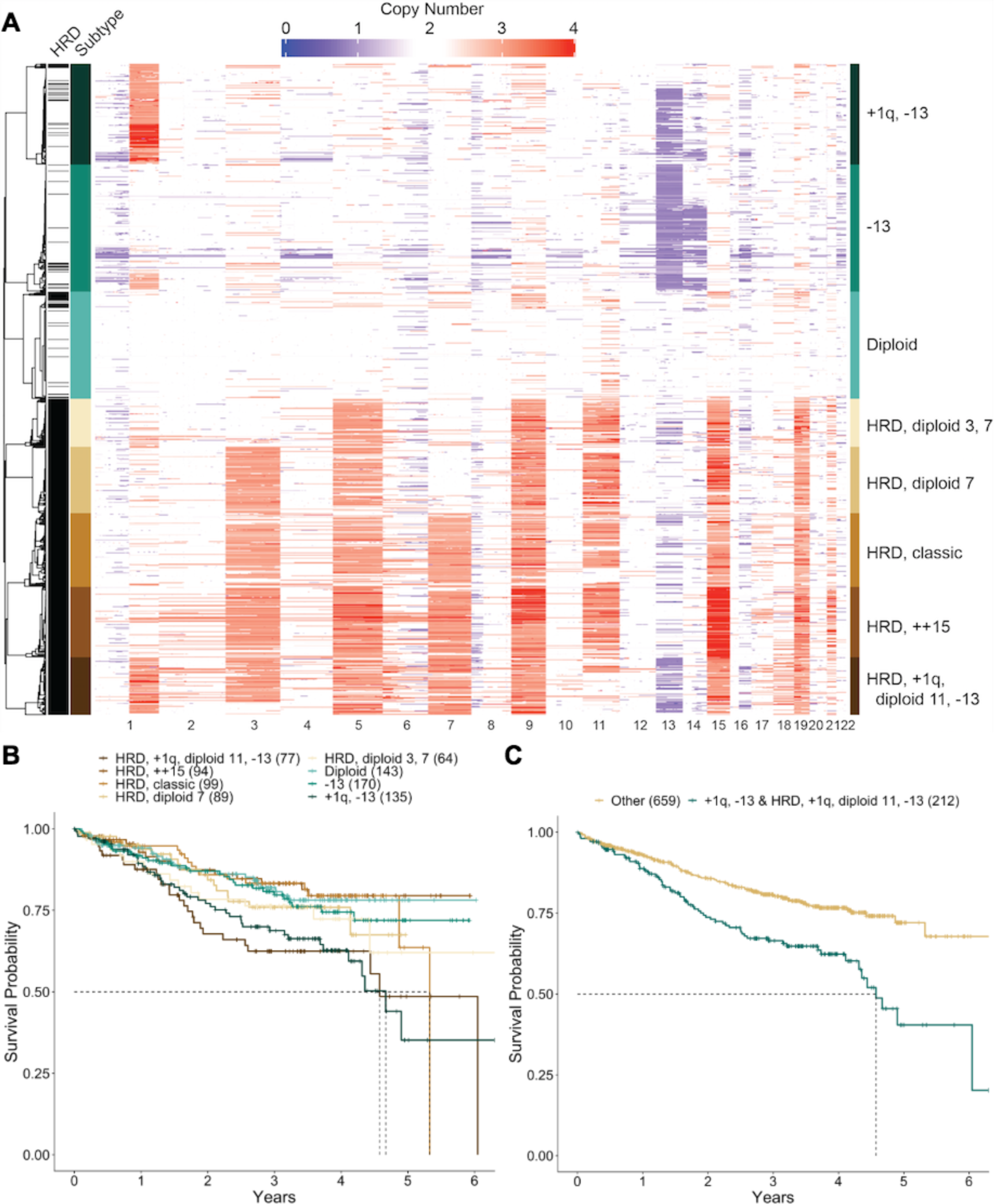
Copy number subtypes of multiple myeloma. (A) Consensus clustering of WGS CN data identified eight unique CN subtypes, comprising five HRD and three NHRD clusters that were annotated based on common CN features. (B) OS outcomes by CN subtype. Median OS was met for the HRD, +1q, diploid 11, -13 (56 months), +1q, -13 (57 months), and HRD, classic (65 months) subtypes. (C) OS outcomes of patients in the +1q, -13 and HRD, +1q, diploid 11, - 13 groups (median = 56 months) versus patients in other CN subtypes (p<0.001).

In the cohort, there was no difference in outcomes between HRD and NHRD patients (Figure S2.5). However, the HRD and NHRD subtypes with both 1q gain and chr13 loss had inferior OS outcomes when compared to patients in other CN subtypes (Figure 2B), suggesting that HRD patients should not universally be considered as a group with good outcomes. Combining these two subtypes identified a group with inferior outcomes as compared to patients with other genetic backgrounds (Figure 2C, HR=1.928, 95% CI=1.435-2.59, p<0.001). The observation that NHRD patients in the +1q, -13 subtype exhibited poor OS outcomes as compared to NHRD patients in the -13 subtype suggested 1q gain, rather than 13q loss, is the predictor of poor outcome, which was confirmed in a cox proportional hazard model examining the contribution of 13q14 and 1q21 CN on OS outcomes across the cohort (Figure S2.6).

### RNA Subtypes of Multiple Myeloma

Consensus clustering was performed on RNA sequencing results from 714 baseline samples to identify subtypes of myeloma defined by gene expression similarities. The number of clusters that best represented the data was 12 across three independent replicates, for which two were identical and a third had 20 (2.8%) patients assigned to different classes (Figure S3.1-3.3). Many of the observed subtypes were associated with known immunoglobulin translocations and CN states (Figures 3A & S3.4) and there were clear relationships with subtypes identified in previous studies (Figures S3.5 & S3.6)^17, 18^. Four subtypes were identified across all studies, including: MS, characterized by t(4;14) patients; MAF, characterized by t(14;16) patients; CD1 characterized by t(11;14) patients; and PR, characterized by patients with a high proliferation index. To maintain consistency across studies we used subtype names from previous studies when appropriate but otherwise assigned names based on common molecular features.

**Figure 3.**
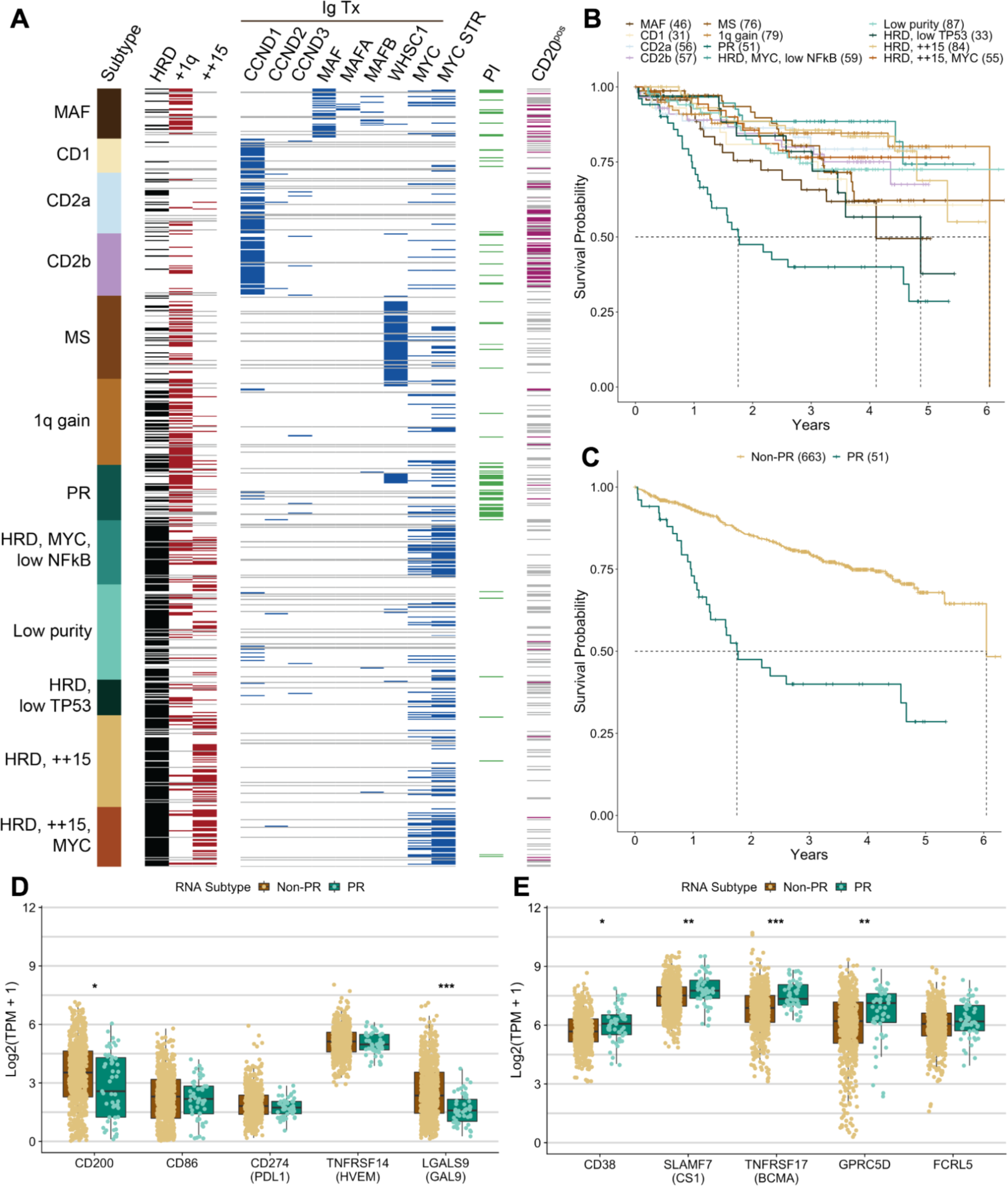
RNA subtypes of multiple myeloma and associated characteristics. (A) Consensus clustering of RNAseq data revealed 12 RNA subtypes of multiple myeloma. (B) OS outcomes for patients by RNA subtype. Median OS was reached for the PR (21 months); MAF (50 months); HRD, low TP53 (59 months); and 1q gain (74 months) subtypes. (C) OS outcomes of patients in the PR versus non-PR (median = 74 months) subtype at diagnosis (p<0.0001, HR = 3.73 [95% CI 2.49-5.58]). Expressed (TPM>1 in at least one group) checkpoint inhibitor (D) and immunotherapy (E) targets in non-PR versus PR patients. A significant difference in median expression between the two groups is designated (***p<0.001, **p<0.01, *p<0.05).

The MS subtype comprised 10.6% of patients for whom a t(4;14)-WHSC1 was detected in 62/67 (92.5%) by WGS. In one patient, a t(2;4) involving the kappa locus was detected. Two of the four remaining patients had detected fusion transcripts between WHSC1 and the highly expressed genes FUT8 or CXCR4. The MAF subtype included 6.4% of patients in which we detected immunoglobulin rearrangements in 38/41 (92.7%) of patients (27 t(14;16)-MAF, 4 t(8;14)-MAFA, 6 t(14;20)-MAFB and 1 t(20;22)-MAFB). All three patients with undetectable immunoglobulin translocations had high expression of a MAF family gene and in two patients the mechanism was identified. One had a t(1;16) juxtaposing the FAM46C super-enhancer with MAF^19, 20^. Another had an atypical insertion of a class-switch circle telomeric of MAF. One patient had both a t(14;16)-MAF and t(4;14)-WHSC1 yet strongly associated with the MAF subtype, suggesting the MAF expression signature can overpower the MS signature. MAF immunoglobulin translocations were associated with higher mutation load^21^ and in this cohort 8/10 patients with high tumor mutation burden (>10 mut/mb) were in the MAF subtype, and could qualify to receive a checkpoint inhibitor.

Three subtypes were highly associated with overexpression of a D-type cyclin caused by t(11;14)-CCND1, t(12;14)-CCND2, or t(6;14)-CCND3 (Figure S3.7A-C). The CD1 subtype included 4.3% of patients 24/25 (96%) had a detected D-type cyclin targeting translocation. The remaining patient had a t(9;14) which linked the immunoglobulin heavy chain locus with the B-cell master regulator PAX5 resulting in its overexpression (Figure S3.7D). Unlike previous studies that identified a single CD2 subtype, we identified two related subtypes designated as CD2a and CD2b. The CD2a subtype comprised 7.8% patients of whom 40/47 (85.1%) had a detected D-type cyclin IgH translocation. The CD2b subtype included 8.0% patients of whom 51/56 (91.1%) had a detected D-type cyclin targeting translocation. Both the CD2a and CD2b subtypes were associated with cell surface expression of CD20, which is largely absent in other RNA subtypes, including CD1.

The PR subtype contained 7.1% of patients with an admixture of classic genetic subtypes and very poor clinical outcome, with a median OS of 21 months (Figure 3B-C). High proliferation index scores were also concentrated in this subtype (Figure S3.8). Clearly, current treatment regimens are ineffective for these patients. We compared the expression of current checkpoint and immunotherapy targets in non-PR versus PR patients and observed that three of five checkpoint targets showed no difference in expression between the two groups, whereas all immunotherapy targets showed either no difference in expression (1 target), or had a higher median expression (4 targets) in PR patients (Figure 3D-E).

A subtype representing 11.1% of patients most closely resembled the previously defined low bone (LB) subtype^17^(Figure S3.5), however, there was no noticeable decrease in bone lesions (Figure S3.9). This subtype comprised an admixture of 59.2% HRD and 40.8% NHRD patients, but 74.0% had a gain of chr1q with 26.0% having ≥4 copies and was thus termed the 1q gain subtype.

Four of the RNA subtypes were associated with a HRD karyotype (Figure S3.4) and either did not uniquely associate with a subtype from a previous study, or the original name could not be justified. Two of the HRD subtypes associated closely with the HY (hyperdiploid) subtype identified previously but differed due to an enrichment of tetrasomy 15, observed in 58.7% and 60.8% of patients. Since structural events involving MYC are associated with HRD karyotypes^22^, we investigated the association between these two groups and MYC rearrangements. We identified 37/49 (75.5%) patients versus 23/76 (30.3%) of patients had MYC rearrangements, and thus these subtypes were named HRD ++15, MYC and HRD ++15 respectively. A third HRD subtype comprising 8.3% of patients most closely associated with the PRL3 subtype^18^, however, the signature was elevated in four subtypes (Figure S3.6). A MYC structural event was identified in 35/49 (71.4%) of these patients and this group was also distinguished from all others except PR in having a low NFkB index (Fig S3.10) and was thus named HRD MYC, low NFkB. The smallest HRD group contained 4.6% of patients and was associated with the previously defined NF-kB subtype^18^, however, no clear association existed with the NF-kB index used to define the subtype (Figures S3.6 & S3.10). One of the predictors of this RNA subtype was overexpression of NINJ1 (Table S7, Figure S3.11A), which inhibits translation of TP53^23^. TP53 was also found to be underexpressed, exhibiting the lowest median expression in this subtype as compared to all other RNA subtypes (Figure S3.11B). Taken together, this subtype was termed HRD, low TP53.

The final subtype contained 12.2% of patients and strongly correlated with the previously defined myeloid group^18^. Analysis of multiple data types indicated a bias to lower purity samples, and was thus termed Low purity (Figures S3.6 & S3.12).

### Clinical and Molecular Associations with RNA Subtypes

Some RNA subtypes were strongly associated with specific molecular events while others seemed to be admixtures with a common transcriptional phenotype. To identify additional defining features of each RNA subtype, we tested for significant associations between clinical and molecular features, including complete LOF and GOF events. Overall, 21 genes with complete LOF or GOF were identified to have a significant association with one or more RNA subtype (Figure 4). As expected, GOF was detected in the translocation target genes associated with the MAF, MS, and the CD subtypes. Although loss of one WWOX allele is expected in t(14;16) we frequently detected complete LOF of WWOX (p<0.001) supporting a possible role of WWOX in myeloma. Both the MS and 1q gain subtypes were diminished for NRAS GOF, and the latter was enriched for TRAF3 LOF. The CD2a subtype was enriched for GOF events in NRAS (p<0.005) and IRF4 (p<0.005) while the CD2b subtype was enriched for GOF events in IRF4 (p<0.005) and EFTUD2 (p<0.01) representing potential subtype-specific therapeutic targets. In general, the HRD RNA subtypes were not enriched for any GOF or LOF events aside from the HRD, ++15, MYC subtype, which was enriched for LOF events in FAM46C (p<0.001).

**Figure 4.**
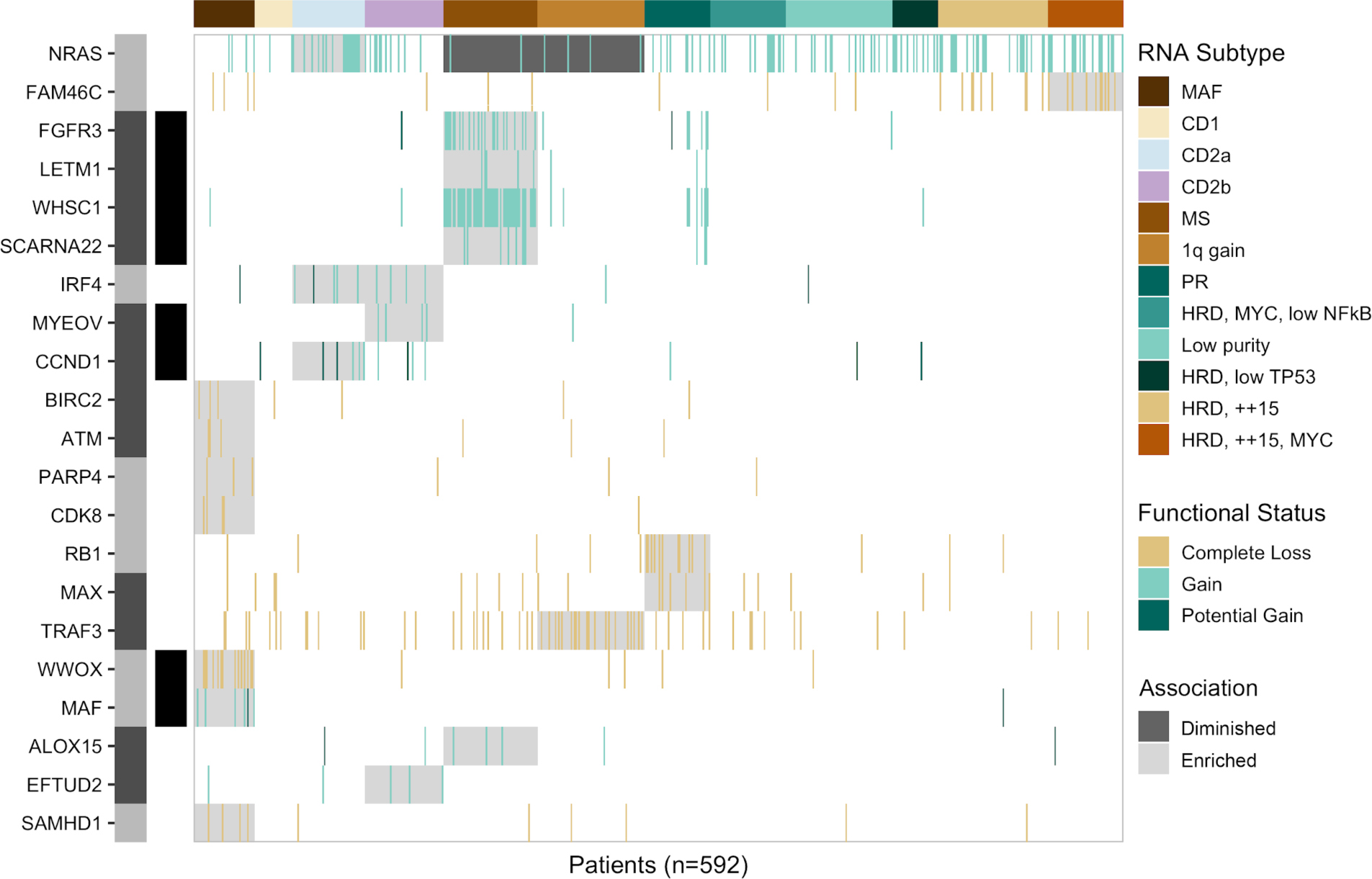
RNA subtypes and significantly associated LOF and GOF events (pFDR≤0.1, p<0.05). The location and proximity of individual genes is shown next to each gene with the alternating gray and black bar illustrating when the chromosomal location changes, while black bars directly to the right denote contiguous genes.

The PR subtype was enriched for LOF of RB1 (p<0.001) and MAX (p<0.01), gain(1q21) (p<0.001), del(13q14) (p<0.001), and ISSIII patients (p<0.001). Interestingly, 50% of PR patients were ISSIII while 22% and 28% were ISSI and ISSII, highlighting that ISS underestimates disease severity in half of these high-risk patients. Different mechanisms of complete loss of RB1 were observed but typically involved a one copy deletion of 13q coupled with a second molecular event (Figure S4.1). Identifying LOF of RB1 and MAX provides the first defining genetic features of the high-risk PR phenotype.

### Transition to PR at Progression and Link with G1/S Checkpoint

To apply the RNA subtype classification to the serial samples, we developed a predictive model that outputs the class probability associated with each of the 12 subtypes and assigned each progression sample the subtype with the highest class probability. Overall 71 patients were assigned a subtype at two or more timepoints, with 55 patients assigned a subtype other than Low purity for at least two time points. At diagnosis, 5 serial patients were classified as Low purity, however, at progression they all had a subtype other than Low purity, further supporting that this phenotype was driven by relative sample purity rather than a distinct disease signature (Figure S5.1). For each patient, subtype assignments were compared across visits. Although most patients remained in the same subtype throughout their disease course 13/51 (25.5%) non-PR patients transitioned into the PR subtype at progression (Figure 5A). Regardless of original subtype, patients that transitioned to the PR subtype rapidly succumbed to their disease (Figure 5B), with a median OS after the detected progression of 88 days (Figure S5.2), and had inferior outcomes compared to other patients who also progressed (Figure 5C).

**Figure 5.**
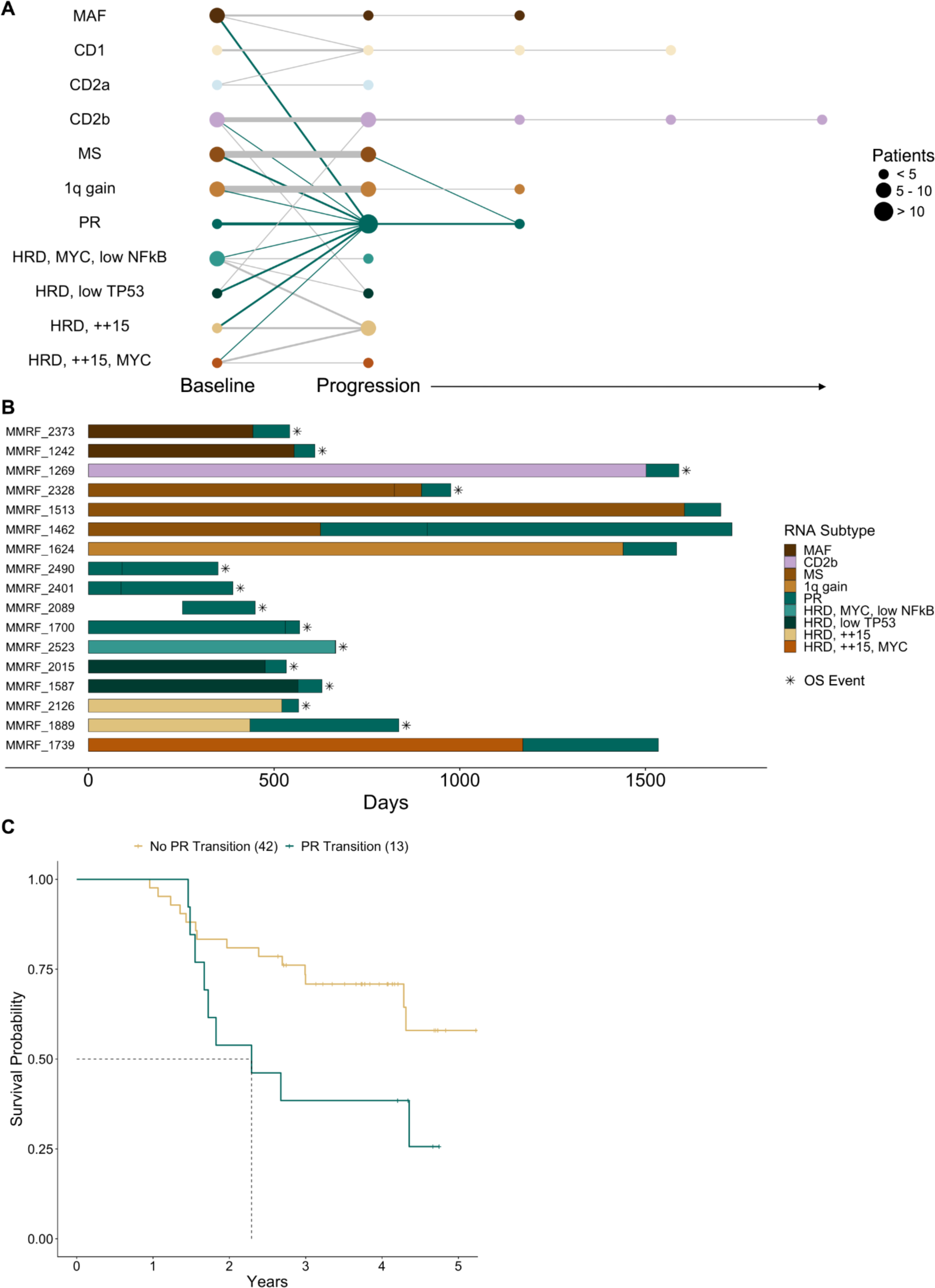
RNA subtype of serial patients at baseline and progression. (A) Node size reflects the relative number of patients in each RNA subtype at each timepoint while edge width reflects the relative number of patients remaining in, or transitioning to, a particular RNA subtype, with the thinnest line and thickest line representing 1 and 7 patients respectively. (B) Swimmers plot of patients in the PR subtype at either baseline or progression. Vertical breaks indicate visits with available RNA sequencing data for RNA subtype prediction. Fill color indicates RNA subtype between visits. Asterisks denote OS events. (C) OS outcomes for serial patients that transition to the PR subtype at progression (median = 28 months) versus those that do not (median not met, p<0.05).

To identify molecular events potentially driving the transition of patients to the PR subtype, gene functional status was compared at the PR and prior non-PR time point. Molecular data was available for comparison at both time points for 9/13 patents that transitioned to PR. Despite the prevalence at baseline, none of the patients transitioning to PR acquired complete LOF of RB1. However, three patients (33%) had complete LOF of a cyclin-dependent kinase inhibitor at progression. Two patients had complete LOF of CDKN2C at progression due to homozygous deletions (Figure S5.3). One patient acquired two independent deletions at progression while the other had one clonal deletion at diagnosis and the second became clonal, increasing from 26% to 100%, at progression, suggesting this aggressive clone existed even before any treatment was received. One patient acquired complete loss of CDKN1B at progression from the combination of a pre-existing deletion and a clonal frameshift mutation detected only at progression, suggesting the mutation either existed at a frequency below our limit of detection, or was acquired (Figure S5.4). Similar to the baseline observations, there are multiple genetic defects in G1/S checkpoint genes that can result in the PR phenotype.

## Discussion

The MMRF CoMMpass study represents the largest sequencing study of multiple myeloma patients undertaken to date based on the number of enrolled patients and the total number of sequencing assays performed. The cohort has facilitated the identification of distinct CN and expression subtypes of myeloma, as well as both recurrent and rare molecular events that occur at frequencies that would not be detected in smaller patient cohorts.

A diverse array of genetic events can contribute to the development or progression of cancer, with individual genes often being affected by multiple types of alterations, however, these diverse processes are generally summarized in isolation. To accurately summarize the frequency of these changes, we integrated seven different data formats extracted from WES, WGS, and RNAseq data and identified 70 LOF and 27 GOF genes occurring in five or more patients. This approach permitted the identification of genes that are often under-represented by a single technology including complete LOF in RB1, CDKN2C, and WWOX, and allowed each gene to be assigned a biologically relevant phenotype of functional, partial loss, or complete loss. Differentiating between partial and complete LOF is pertinent for accurate identification of high-risk patient populations. In TP53, solitary deletions or mutations have been associated with poor prognosis, however, only patients with complete LOF of TP53 have poor outcomes, suggesting only one third of patients with del(17p13) identified by clinical cytogenetic assays are true high- risk patients^12, 24–2626^. Finally, there is a long standing interest in determining the gene associated with monosomy 13 in myeloma. Our analysis identified recurrent complete loss events in RB1 and DIS3, but also identified independent complete loss events in PSPC1, TGDS, LATS2, BRCA2, PARP4, MYCBP2, TPP2, CDK8, TSC22D1, and ARHGEF7 suggesting multiple genes on chr13 can independently contribute to myelomagenesis.

Few studies have performed CN clustering and, due to their limited sample size, have barely resolved distinct CN subtypes beyond the classic HRD and NHRD phenotypes^3^. We identified eight distinct CN subtypes, including five HRD, and three NHRD subtypes. Although previous studies have shown HRD patients have favorable outcomes compared to NHRD patients, in CoMMpass there was no difference in OS or PFS outcomes. This large cohort analysis revealed a number of seemingly inter-related events such as 1q gains and monosomy 13; in HRD patients lacking trisomy 3, a concomitant absence of trisomy 7; and groups with a classic HRD phenotype defined by trisomy or tetrasomy 15. Interestingly, there were independent HRD and NHRD subtypes with 1q gain and chr13 loss with the HRD group lacking the classic trisomy 11, suggesting the combination of these events can phenocopy the benefits of trisomy 11. Although patients with 1q gain and 13 loss represent poor outcome CN subtypes, the median OS of these patients was just under 5 years, comparable to the 4.5 year OS associated with R-ISSIII which includes patients with high-risk clinical and cytogenetic features^15^. Taken together, this highlights that CN features alone are insufficient to distinguish the subset of ultra high-risk myeloma patients with early OS events.

Previous studies clustering myeloma gene expression data have identified 8 to 10 unique subtypes, many of which were consistent among studies including the MS, MAF, CD1, CD2, and PR subtypes^17, 18^. Our consensus clustering of RNAseq data from CoMMpass identified 12 unique RNA subtypes. In this study, some previously identified subtypes were further broken down, while others were renamed to better reflect the underlying biology, which was aided by incorporating the multitude of data types in this study. For example, previous studies identified a single CD2 group, however, we identified two CD2 groups, designated CD2a and CD2b, which were associated with IRF4 mutations, PAX5 expression, and CD20 surface expression. Several studies have sought to identify treatment strategies for CD20 positive patients, but based on the distribution within CD2a and CD2b it may be pertinent to consider that these patients originate from two unique populations. In the context of precision medicine, the strong link between t(11;14) and the CD subtypes leads to question whether one of the CD subtypes better predicts response to venetoclax than t(11;14)^27^.

The PR RNA subtype defined a group of patients with extremely poor OS, high proliferative index scores, and nearly ubiquitous 1q gains. This group has remained controversial because of the competing supervised patient segregation TC classification models^28, 29^, that argue to group patients by defined genetic features, however, given the mixture of genetic backgrounds observed in this subtype, it would be difficult to identify these high-risk patients based on genomic features alone. Through our integrated analysis we identified LOF of RB1 or MAX as common genetic events in baseline PR patients, which provides the first genomic link to this gene expression phenotype. Loss of MAX was recently associated with transformation and increased proliferation in small cell lung cancer^30^ and thus, LOF of RB1 or MAX likely contributes to the highly proliferative phenotype observed in PR patients. PR patients exhibited similar or lower median expression of checkpoint targets but higher expression of most immunotherapy targets when compared to non-PR patients suggesting that immunotherapies may represent a viable therapeutic option in these high-risk patients, and highlighting the importance of identifying these patients in future clinical studies.

There was also a strong tendency for patients to transition to the PR subtype at progression, with 25.5% of serial patients in a non-PR subtype at diagnosis transitioning to PR. Patients that transitioned to the PR subtype had extremely poor outcomes after the transition, with a median survival of less than three months after their progression visit. An acquired complete loss of a cyclin-dependent kinase inhibitor, such as CDKN2C or CDKN1C, was observed in 33% of patients transitioning to the PR subtype at progression suggesting that transition to the PR phenotype at progression is highly associated with genetic events that further disrupt cell cycle control.

These findings demonstrate that advanced molecular diagnostics such as WGS and RNAseq are better predictors of disease behavior than current staging systems based on clinical laboratory, conventional cytogenetic, and FISH data. These assays also identify therapeutic targets including RAS, BRAF^V600E^, and FGFR3 mutations, that are actionable with agents already approved for other cancer indications. In fact CoMMpass findings drove the development of a number of clinical trials, notably, MMRC-085 Myeloma-Developing Regimens Using Genomics (MyDRUG, NCT03732703), an umbrella trial using targeted exome and transcriptome sequencing to stratify subjects into sub-protocols with approved targeted agents. These findings favor adoption of advanced molecular diagnostics into the routine care of myeloma, especially as the breadth of clinical impact broadens and costs decrease.

Comprehensive molecular analyses of the baseline CoMMpass cohort has permitted a more thorough understanding of the genetic diversity and subtypes of the disease. This approach clearly defined the primary molecular features driving different subtypes of multiple myeloma and identified high-risk patients at both diagnosis and progression. Innovative clinical trials targeting this high-risk population are needed given the current poor outcomes with therapies that are otherwise highly effective in other subtypes. Given that patients frequently transition to the high-risk PR subtype at progression, it will be important to know the percentage of PR patients in clinical trial populations, particularly in the relapse/refractory setting, to understand if the arms are balanced and if there is a difference in response between these groups. The identification of unique subtypes and the frequency of target gene dysregulation, via our integrated analysis, provides a solid foundation to prioritize targets for precision medicine approaches in multiple myeloma.

## Methods

### Sample Collection and Biobanking

All samples and clinical data analyzed in this study were collected as of interim analysis 14 from patients who consented to participate in the Relating Clinical Outcomes in Multiple Myeloma to Personal Assessment of Genetic Profile (CoMMpass) Study (NCT01454297), sponsored by the Multiple Myeloma Research Foundation (MMRF). The MMRF CoMMpass study accrued patients from clinical sites in Canada, Italy, Spain and the United States. All patient samples were shipped to one of three biobanking operations: Van Andel Research Institute (VARI) in Grand Rapids, Michigan for all samples collected in Canada or the United States; University Hospital of Salamanca for samples collected in Spain; or University of Torino for samples collected in Italy. In North America and Spain, K2-EDTA tubes were used for collection of peripheral blood (PB) and sodium heparin tubes were used for the collection of bone marrow (BM) aspirates. These samples were shipped to their respective biobank using CoMMpass study kits that maintained samples at 7-12°C. In Italy, clinical sites participating in the FORTE clinical trial (NCT02203643) collected BM and PB sample aspirates in sodium citrate vacutainers. Samples collected at sites in Italy were shipped at ambient temperature to the biorepository site.

At VARI, the received BM and PB specimens were first quality controlled by flow cytometry to determine the percentage of plasma cells (PCs) in the PB and BM specimens. Patients were only included in the study when the submitted BM contained at least 1% PCs. If the PB showed less than 1% circulating PCs, white blood cells were used as the constitutional DNA source, however, when >1% circulating PCs were observed enriched CD3 positive T-cells were used. Whole BM PC enrichment, or PB PC enrichment when >5% circulating PCs were detected, was performed using the Miltenyi autoMACS Pro Separator using anti-CD138 microbeads. The purity of the enriched samples was assessed using a 3-color slide-based immunofluorescence assay that identified cells with DAPI and the presence or absence of kappa or lambda light chains. Clinically eligible baseline patients with tumor samples with greater than 250,000 cells recovered after CD138 enrichment and monoclonal purity greater than or equal to 80% moved forward for nucleic acid extraction. For progression samples the cell recovery requirement was 200,000 cells. To minimize nucleic acid isolation failures, the first 750,000 cells were used exclusively for DNA isolation. When more than 750,000 cells were recovered, the sample was split 50/50 for DNA and RNA isolations. When more than 4 million cells were recovered, multiple aliquots were stored for future use. Cells destined for DNA isolation were stored as snap frozen pellets, while samples for RNA extraction were lysed in QIAzol before long-term storage at -80°C.

Samples from clinical sites in Spain were collected and shipped using the provided CoMMpass collection kits. Red blood cells were removed from the PB and BM specimens using a red cell lysis buffer and, following a PBS wash, the remaining white blood cells were counted. After red cell lysis, the isolated cells were quality controlled using flow cytometry to determine the percentage of PCs in the PB and BM specimens. If the PB showed less than 1% circulating PCs, white blood cells were used as the constitutional DNA source; however, when >1% circulating PCs were observed, enriched CD3 positive T-cells were used. The isolated PB cells (1-5 million cells) were snap frozen as dry pellets for constitutional DNA isolation. The isolated BM cells were stained with anti-CD138 microbeads and PCs were enriched using a Miltenyi autoMACS Pro Separator. The enriched CD138^+^ cells were stored as snap frozen dry pellets and, when possible, a separate aliquot was lysed with QIAzol and stored at -80°C until shipped on dry ice to VARI for isolation. The purity of the CD138 enriched PC fractions was determined using flow cytometry with antibodies against CD38, CD138, and CD45.

Samples collected in Italy were treated with red cell lysis buffer. After washing the remaining WBC with PBS, the cells were counted. The isolated PB cells (1-5 million cells) were snap frozen as dry pellets for constitutional DNA isolation. The BM WBC were stained with anti- CD38 magnetic beads and PCs were enriched using a Miltenyi autoMACS Pro Separator. After sorting, the purity of the enriched specimens was assessed by flow cytometry using a fluorescent anti-CD38 antibody. Cells destined for DNA isolation were stored as snap frozen pellets, while samples for RNA extraction were lysed in QIAzol before long-term storage at -80°C.

### Flow Cytometry Phenotyping and Quality Control Process

All samples received at VARI were tested by flow cytometry to phenotype and quality control the received specimens. The antigens and corresponding commercial antibodies used in the flow cytometry assays are as follows: CD38 (BD Biosciences, 340677), CD45/PTPRC (BD Biosciences, 340665), CD138/SDC1 (BD Biosciences, 347205), CD319/SLAMF7 (Invitrogen/eBioscience, 12-2229-42), CD13/ANPEP (BD Biosciences, 340686), CD19 (BD Biosciences, 340720), CD20/MS4A1 (BD Biosciences, 346581), CD27 (BD Biosciences, 654665), CD28 (BD Biosciences, 348047), CD33 (BD Biosciences, 340679), CD52 (Life Technologies, MHCD5204), CD56/NCAM1 (BD Biosciences, 340724), CD117/KIT (BD Biosciences, 340867), FGFR3/CD333 (R&D Systems, FAB766P), Kappa (BD Biosciences, 643774), and Lambda (Life Technologies, MH10614). Flow panels performed included CD38 x CD45 x CD138 x CD56 (initial BM & PB screening panel 1); CD38 x CD45 x CD138 x CD319 (updated screening panel 1 after the introduction of daratumumab); CD38 x CD45 x cytoplasmic Kappa x cytoplasmic lambda (BM & PB screening panel 2); CD38 x CD45 x CD138 x (either CD13, CD19, CD20, CD27, CD28, CD33, CD52, CD117, FGFR3); and propidium iodide stained nuclei to determine the DNA content of each tumor.

### Nucleic Acid Isolation

All nucleic acid isolations were performed at VARI. DNA was extracted from the dry cell pellets with the Qiagen Gentra Puregene Tissue Kit (Qiagen, 158667) with isolated DNA suspended in Qiagen buffer ATE, and stored at -20°C. DNA was extracted from PB samples using the Qiagen QIAsymphony, which uses magnetic beads for automated sample processing. Blood tubes were either processed immediately upon receipt, or frozen at -20°C and processed in batches. QIAsymphony extractions were performed using the DSP DNA Midi Kit (Qiagen, 937255). DNA was eluted in Qiagen buffer ATE and stored at -20°C. DNA was quantified by Nanodrop spectrophotometric analysis, as well as by fluorescence using the Qubit 2.0 to determine dsDNA content. Sample quality was determined by agarose gel or Agilent TapeStation Genomic ScreenTape. Samples with at least 250 ng of dsDNA were submitted to TGen for analysis.

Tumor cells designated for RNA extraction were dissolved in QIAzol Lysis Reagent (Qiagen, 79306), stored at -80°C, and extracted with the Qiagen RNeasy Plus Universal Mini Kit (Qiagen, 73404). RNA was eluted in nuclease-free water and stored at -80°C. RNA was quantified by Nanodrop spectrophotometric analysis and RNA quality was evaluated using the Agilent Bioanalyzer 2100. Samples with a RIN ≥6.0 and at least 200 ng of RNA were submitted to TGen for analysis.

### RNA Sequencing (RNAseq) Library Preparation

All RNA sequencing libraries were constructed using the Illumina TruSeq RNA library kit following the manufacturer’s recommendations. Over the course of the project the primary target input changed from 2000 ng to 500 ng. When 500 ng of RNA was not available, samples were processed using a lower input of 250 ng or 150 ng. In all cases, the input quantity was based on nanodrop quantification. RNA quality was confirmed at TGen to have a required RIN ≥ 7 using an Agilent Bioanalyzer with the RNA 6000 Nano Kit (Agilent, 5067-1511) or eRIN ≥ 6 on an Agilent Tapestation using the RNA ScreenTape assay (Agilent, 5067-5576, 5067-5577). Libraries were prepared following the manufacturer’s recommendation for the TruSeq RNA Library Prep Kit v2 (Illumina, RS-122-2001), unless otherwise noted. Eight cycles of PCR amplification for 2000 ng and 500 ng input, 9 cycles for 250 ng input, and 10 cycles for 150 ng input were performed. Final libraries were quantified using the Qubit dsDNA HS Assay Kit (Invitrogen, Q32854) and assessed on the Agilent TapeStation using the High Sensitivity D1000 ScreenTape assay (Agilent, 5067- 5584, 5067-5585).

### Long Insert Whole Genome Sequencing (LI-WGS) Library Preparation

LI-WGS libraries were initially generated using the Illumina TruSeqDNA Whole Genome kit (TSWGL). Fragmentation of 600-1100 ng of DNA was performed on a Covaris E210 Focused Ultrasonicator with the following parameters: duty cycle = 2, peak power = 6, cycles per burst = 200, and time = 20 sec. Libraries were prepared according to the standard Illumina protocol, however, the AMPure bead ratios were adjusted to either 1:1 or 1:0.8. After ligation clean up, samples were run on a 1.5% agarose gel and Xtracta gel extractors (USA Scientific, 5454-2500) were used to extract library molecules at approximately 1.3, 1.0, and 0.8 kb. Size-selected samples were processed with Freeze ‘N Squeeze DNA Gel Extraction Spin Columns (Bio-Rad, 732-6165). Purified 1 kb samples were concentrated using AMPure XP beads and amplified with 10 PCR cycles. Final libraries were run on Agilent Bioanalyzer DNA 12000 chips (Agilent, 5067- 1508 & 6067-1509). If a patient’s tumor and constitutional samples were not within ∼100 bp of each other, alternate excised samples (1.3kb or 0.8kb) were processed to generate a better match.

For samples processed using the KAPA HTP/LTP Library Preparation Kit (Roche, KK8234) and off-bead protocol (KAWGL), 200-1100 ng of DNA was fragmented on the Covaris E210 (see parameters above) or E220 with the following parameters: duty cycle = 4%, initial / peak power = 170 W, cycles per burst = 200, and time = 20 sec. Libraries were prepared according to the standard Kapa protocol, however, the AMPure bead ratio was adjusted to either 1:1 or 1:0.8. Libraries were amplified for two cycles before size selection to linearize the fragments and five cycles after size selection using KAPA HiFi HotStart ReadyMix (Roche, KK2602) For samples processed using the KAPA HyperPrep kit (Roche, KK8505) (KHWGL), 200 ng of DNA was fragmented on the E210 or E220 Covaris with the parameters defined above. Libraries were prepared according to the manufacturer’s protocol, however the post ligation AMPure bead ratio was adjusted to 1:0.8. Molecules between 950-1050 bp were either extracted automatically from a Sage Science Pippin Prep 1.5% gel (Sage Science, CSD1510) or hand punched from a 1.5% agarose gel. One cycle of PCR amplification pre size selection, followed by 6 cycles of amplification post size selection, was performed using KAPA HiFi HotStart ReadyMix.

### Whole Exome Sequencing (WES) Library Preparation

Initially samples were processed using the Illumina TruSeq Exome Library Prep Kit (TSE61) until the product was discontinued. Genome libraries were created using 1100 ng of DNA as input for fragmentation on the E210 Covaris using the following settings: duty cycle = 10, intensity = 5, cycles per burst = 200, time = 120 seconds. Individual DNA samples were mixed with TElowE (10 mM Tris-HCL, 0.1 mM EDTA) to a final volume of 55 μl in a covaris microTUBE plate. Six libraries were pooled together before enrichment following the manufacturer’s protocol.

Subsequently, samples were processed using the KAPA HTP/LTP Library Preparation Kit using the off-bead protocol and 8-plex pooled enrichment was performed using Agilent SureSelect XT2 Human All Exon V5+UTR baits (Agilent, G9661B) (KAS5U). Genome library preparation was performed according to the manufacturer’s protocol with 500 ng of input DNA fragmented to an average target size of 160 bp using a Covaris E220 focused-ultrasonicator with the following settings: duty cycle 10%; peak power 175; cycles per burst 200; time 300; power mode “frequency sweeping”; bath temperature 7°C. Five cycles of PCR amplification was performed pre-capture using KAPA HiFi HotStart ReadyMix, and the resulting libraries were quantified using either the Agilent Bioanalyzer 1000 chip (Agilent, #5067-1504) or the Agilent Tapestation D1000 kit (Agilent, 5067-5582, 5067-5583). Eight libraries were pooled at 187.5 ng each before capture following the Agilent XT2 protocol. The enriched library pool was amplified for 8 cycles using KAPA HiFi HotStart ReadyMix. Final libraries were quantified using the Agilent Bioanalyzer High Sensitivity DNA Kit (Agilent, #5067-4626) or Agilent Tapestation HSD1000 (Agilent, #5067- 5584 & 5067- 5585) and Qubit High-Sensitivity assay (Invitrogen, #Q32854). Samples processed using the KAPA HTP/LTP Library Preparation Kit and the on-bead protocol followed by 8-plex pooled enrichment performed using Agilent SureSelect XT2 Human All Exon V5+UTR baits (KBS5U) were performed according to the manufacturer’s protocol with 500 ng of input DNA as described above for the KAS5U protocol.

Samples processed using KAPA HyperPrep Kits were prepared on an Agilent Bravo liquid handler followed by single-plex Agilent SureSelect XT enrichment using V5+UTR baits (KHS5U). Genome libraries were prepared from 200ng, 100ng, or 50ng of dsDNA. Each sample was fragmented to an average target size of 160 bp using a Covaris E220 with the following settings: duty cycle = 10%; peak power = 175; cycles per burst = 200; time = 300; power mode = “frequency sweeping”; bath temperature = 7°C. Pre-capture PCR using 7, 8, or 9 cycles of amplification was performed for the 200, 100, and 50 ng inputs, respectively, followed by 8 cycles of post-capture PCR.

### Illumina Sequencing

Sequencing was performed on an Illumina HiSeq2000 or HiSeq2500 at TGen using Illumina HiSeq v3 or v4 chemistry. Diluted library pools with 1% PhiX control libraries were clustered on Illumina cBOT instruments as recommended by the manufacturer. In all cases, sequencing assays used a paired-end sequencing format with at least 82x82 nucleotide reads. The majority of RNA sequencing libraries, which all had 6 bp sample indexes, were sequenced using paired-end 83x83 reads. Exome libraries with 6 bp sample indexes were sequenced using paired-end 83x83 reads, while those with 8 bp sample indexes were sequenced using 82x82 reads. Whole genome long-inserts were typically clustered on individual lanes, allowing a paired-end 86x86 format. In all cases, raw sequencing data was extracted from the BCL files in the resulting Illumina run folders using BCL2FASTQ v1.8.4 or BCL2FASTQ v2.17.1 to generate industry standard FASTQ files.

### Sequencing Data Analysis

Analysis of all sequencing data was performed at TGen on a high-performance computing system using an internally developed analysis pipeline (Medusa Subversion, https://github.com/tgen/medusaPipe) and the MMRF CoMMpass specific TGen05 recipe. This recipe is based on the hs37d5 version of the GRCh37 reference genome used by the 1000 genomes project, with gene and transcript models from Ensembl v74. Additional automated CoMMpass specific primary processing was also performed (https://github.com/tgen/Post_Medusa_Processing). Code for the creation of the reference genome and gene models used, as well as secondary and tertiary analysis methods, are available on GitHub (https://github.com/tgen/MMRF_CoMMpass).

The paired-end fastq files generated in the LI-WGS and WES assays from each sequencing lane were aligned with bwa (v0.7.8-r455). The output SAM file was converted to a BAM file and sorted using SAMtools (v0.1.19-44428cd), after which base recalibration was performed using GATK (3.1-1-g07a4bf8). When multiple lanes existed they were merged into a single BAM file and duplicate reads were marked using Picard (v1.111(1901)), and joint indel realignment was performed using GATK to produce the final BAM files used for analysis. The quality of each assay was determined using multiple Picard and Samtools quality control metrics. To be included in the analysis, both the tumor and constitutional sample needed to meet the following criteria for genomes: physical coverage ≥25x, chimera read rate <3%, and dlrs ≤0.2; and for exomes: >90% target bases at 20x coverage, and chimera read rate <3%. Somatic mutations were identified using Seurat (v2.6, https://github.com/tgen/seurat), Strelka (v1.0.13), and MuTect (v2.2-25-g2a68eab), and their outputs were combined to identify somatic events called by at least two callers. The coding effect of each mutation was determined using snpEFF (v4.2 (build 2015-12-05)), and additional annotations were added using snpSIFT. Somatic structural abnormalities were detected using Delly (v0.7.6) to which additional filtering fields were added to ensure informative read pairs spanned at least a 100 bp window on both breakends. Somatic copy number (CN) abnormalities were identified with a CoMMpass specific implementation of tCoNut (https://github.com/tgen/MMRF_CoMMpass).

Paired-end fastq files from the RNA sequencing assays were aligned using STAR (v2.3.1z 01/24/2013) and the output SAM file was converted to a BAM file and sorted using samTools followed by duplicate marking with Picard. For RNAseq to be included in the analyses, we required at least 50 million read-pairs (100 million reads) generated from each library, a 5’ bias ratio ≥0.5, and a 5’/3’ bias ratio ≥0.75. Gene expression estimates were determined using multiple tools. Counts were extracted from the unsorted SAM file using HtSeq (v0.6.0). TPMs were estimated with Salmon (0.7.2) using the fastq reads as input for quasi-alignment to a transcriptome defined by the GTF gene model. To correct for the variable level of immunoglobulin transcription between samples that compress the TPM values we removed plasma-cell specific transcripts including immunoglobulin, mitochondrial, rRNA, and chrY genes from the final TPM calculation (https://github.com/tgen/Post_Medusa_Processing). Gene fusion events were identified using the TopHat-Fusion workflow in TopHat2 (2.0.8b) followed by independent cross-validation that an associated genomic event existed in the matched LI-WGS assay.

The genotypes of each result file were compared using SNPsniffer (v5) to ensure the constitutional DNA, tumor DNA, and tumor RNA were from the same individual and that each patient was uniquely represented. To ensure accurate alignment with clinical data, molecular predicted gender was required to match the clinically recorded gender and, when available, the clinical immunoglobulin isotype was confirmed to match the isotype defined by flow cytometry and RNA sequencing. All constitutional DNA samples were manually reviewed to ensure they represented a diploid genome. Potential low level cross contamination of the tumor DNA specimens was determined by comparing the number of high confidence mutations detected versus the percentage of those mutations which exist in dbSNP.

### Survival Analyses

Survival curves were computed using the Kaplan-Meier method as implemented in R by the survfit function from the survival (v3.1-8) package and plotted using the ggsurvplot function from the survminer (v0.4.6) package. Pairwise comparisons of survival curves were calculated using pairwise_survdiff from the survminer package. Progression-free survival estimates were computed using the PFS_Censor_Flag (inv_censpfs) and Time_To_Censored_PFS (inv_ttcpfs) fields (Table S1). Overall survival estimates were computed using OS_Censor_Flag (censos) and Time_To_Censored_OS (ttcos) fields. Univariate and multivariate Cox proportional hazards models were calculated using the coxph function from the survival package.

### Integrated Analysis for Gain and Loss of Function Genes

To predict the functional status of each gene in each sample, we performed an analysis integrating multiple measurements from different sequencing assays for each sample. The functional state of each gene was predicted independently to capture loss-of-function (LOF) and gain-of-function (GOF) events. This analysis leveraged 7 outputs from WGS, WES, and RNAseq data for samples with all three data types (592 baseline samples). We sourced the somatic gene level CN and structural abnormalities (deletion, inversion, duplication, translocation) from WGS; the somatic non-synonymous (NS) mutations, B-allele frequency (BAF) and constitutional loss of function mutations from WES; and the gene expression (TPM), cohort normalized median absolute deviation (MAD) expression, and inframe fusion transcripts from RNAseq.

The gene model from Ensembl v74 defines 63,677 distinct genes or gene elements, however, only 57,997 of these map to a contig defined by the hs37d5 reference genome used in this study, and only 57,736 of these map to a primary contig (chromosomes 1-22, X, and Y). The list of genes was reduced to a final list of 23,221 analyzed genes by excluding immunoglobulin elements, excluding genes whose gene names were missing or contained “.” or “-”, and requiring the genes were annotated with the following biotypes: lincRNA, miRNA, processed_transcript, protein_coding, snoRNA, snRNA, TR_C_gene, TR_D_gene, TR_J_gene, TR_J_pseudogene, TR_V_gene, TR_V_pseudogene.

### Loss-of-Function (LOF)

For the LOF analysis we limited the genes to potential tumor suppressor genes (TSG) based on the assumption that these genes would be detectably expressed in the majority of the cohort. To perform this filtering step, 71 known TSGs were obtained from the Cosmic Cancer Gene Census (v75) (http://cancer.sanger.ac.uk/census) that existed in our gene model. For a gene to be included in the LOF analysis, the median expression of the gene in the baseline CoMMpass cohort had to be greater than the median of the 10^th^ percentile of the gene expression of all 71 TSGs. A total of 10,577 genes were included in the LOF analysis after applying the above filter.

In the LOF analysis, a gene was defined as being functional, or exhibiting complete or partial LOF. Complete LOF implied that all functional alleles of the gene are lost, whereas partial LOF implied that there is evidence of inactivation of one allele, and a potential haploinsufficiency. For example, a somatic deletion, NS mutation, structural rearrangement within a gene body, copy neutral LOH, or a constitutional LOF mutation would result in partial LOF of a gene, whereas two or more of these events impacting a gene would result in complete LOF. We used a heuristic approach to determine the functional state of a gene where each position of an 11 bitset identified the presence or absence of a specific genetic event category. The bits and their corresponding definitions in order are: Cl, 1-copy loss; Cd, 2-copy loss; Cn, copy neutral (2-4 copies); M2, two or more NS mutations; M1, one NS mutation; Lh, loss-of-heterozygosity; Ne, gene not expressed; sD, structural deletion; sT, structural translocation; sI, structural inversion; Gm, constitutional SNP. These 11 bits were summarized to assign each gene a status of complete loss, partial loss, or functional.

The segmented CN data was transformed into a sample by gene matrix where each entry represented the lowest log2 fold change observed overlapping the gene body (Table S5). These entries were translated into three flags: ‘*Cd’* for homozygous deletions or 2-copy loss; ‘*Cl’* for 1-copy or heterozygous loss; and ‘*Cn’* designating 2-4 gene copies. We used a log2 threshold of ≤- 2.32 to identify homozygous deletions, which represents the theoretical value if a homozygous deletion exists in 80% of cells, while the remaining 20% of cells are in the normal copy state. A threshold of ≤-0.1613 was used to identify single copy heterozygous losses, which represents the theoretical log2 value if a single copy deletion exists in ∼21.1% of cells.

The only somatic mutations included in the analysis were those annotated as NS mutations with the most damaging effect of the mutation to any transcript being the source of the effect used in the LOF model (Table S2). The count of these mutations per gene was used to define the ‘M1’ bit, which is set when a single mutation is observed, and the ‘M2’ bit when multiple NS mutations were detected. A single NS mutation in a gene, ‘M1’=1, was considered a partial loss and could contribute to complete loss if an additional event from any other mechanism (ie. CN loss) was also observed. When two independent NS mutations were detected in a gene, ‘M2’=1, we assumed they exist in trans, which would represent a bi-allelic loss and thus could be interpreted as a complete LOF based on mutation data alone.

To identify regions of copy neutral LOH we integrated the observed copy number and B-allele frequency (BAF). To set the ‘*Lh’* bit, we extracted the largest overlapping segment from the absolute BAF segmentation data, and set ‘*Lh’=1* when ‘*Cn’=1* and the absolute BAF value was between 0.45 and the max value of 0.5 (Table S5).

To capture loss of gene expression through potential secondary mechanisms, such as epigenetic repression, we identified genes with outlier loss of expression by integrating MAD analysis and the absolute expression value. We used the MAD approach utilized for Cancer Outlier Profile Analysis (COPA) to normalize the gene expression TPM values ^32^. Using this approach, the TPM for each gene was median centered and scaled to their median absolute deviation (MAD). Given the TPM of a gene, the MAD value was computed as:

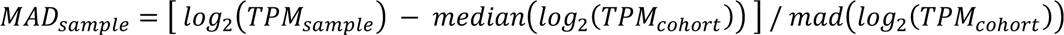

A gene was defined as not expressed if the MAD value was less than the 25th percentile value minus 1.5 times the interquartile range (IQR) and the observed corrected TPM was less than 0.1. When these criteria exist the *‘Ne’* bit was set to 1 to denote a potential epigenetic loss of RNA expression.

To identify when an allele of a gene was inactivated by a structural event we annotated each breakend of structural abnormalities detected by Delly (deletions, inversions, duplications, translocations) independently with an intersecting gene if the breakend occurred between the start and end positions of the gene. To minimize potential double counting of deletions detected by copy number analysis and structural analysis, we only included deletions smaller than 15kb, as the copy number analysis rarely detected deletions below 20kb. When an intersecting gene was annotated on an individual translocation, deletion, or inversion breakend we set the ‘sT’, ‘sD’ and ‘sI’ bits, respectively.

Individuals can also inherit a defective copy of a gene contributing to a LOF event when the remaining functional allele is lost. To identify inherited LOF events we performed joint variant calling on the constitutional exomes from the entire MMRF cohort using GATK (v3.5) best practice guidelines. Individual gVCFs were created using HaplotypeCaller with -L to limit the analysis to the regions targeted by the exome kit. The patient specific gVCF files were combined in batches of 100 using CombineGVCFs and then joint SNV and indel calling was performed across the batched gVCFs using GenotypeGVCFs. Variant quality recalibration (VQSR) was applied independently on the SNP and INDEL detected to reduce false positives. While applying the recalibration, any SNP below the 99.0 tranche and INDEL below the 95.0 tranche were marked as a PASS. These variants were annotated using GATK VariantAnnotator to flag when a variant existed in one of the following databases: dbSNP (v147), ExAC (v3.0), ClinVar (v20170530), NHLBI (v2 ssa137), 1000 Genomes (Phase 3), and COSMIC (v78). To identify highly damaging LOF variants we annotated the variants independently using snpEFF (v4.2) and VEP(v87)/LOFTEE following established rules^33^. Expected LOF variants were extracted from the annotated files when a heterozygous variant was marked as LOF by snpEFF or a high-confidence LOF by LOFTEE and the allele frequency in the CoMMpass cohort was less than or equal to 0.05, and the variant was marked as PASS or the variant was known in dbSNP or ExAC. Variants annotated as LOF by both tools were then annotated with the observed allele ratio in the matching tumor exome and tumor RNAseq alignments. Using the available data, the *‘Gm’* bit was set to indicate if the inherited LOF variant was detectable in the tumor at an expected allele frequency. In the absence of other somatic events, the bit was set when the AR was at least 0.4. When a CN loss was detected we required the AR to be above 0.6 to confirm the LOF impacted the remaining allele, and when copy neutral LOH was detected we required the AR to exceed 0.85.

After setting each bit for each gene in every sample we used a simple heuristics approach to combine and score these events to assign each gene an interpreted functional state. Each bit was assigned a specific weight when set (Cl=0.5, Cd=1.0, Cn=0, M2=1.0, M1=0.5, Lh=0.5, Ne=0.5, sD=0.5, sT=0.5, sI=0.5, Gm=0.5) and these weights are summed to get a final LOF score where values ≥ 1 denote complete LOF, values = 0.5 denote a partial LOF (haplo-insufficiency), and values = 0 denote genes that are expected to be fully functional.

### Gain-of-Function (GOF)

To identify potential oncogenes, we performed a GOF analysis designed to detect common oncogene activation mechanisms including activating mutations, gene amplification, RNA over- expression with concomitant structural events, and inframe gene fusions. The status of each gene was summarized into a 6 bitset which identified the presence or absence of a specific genetic event. The bits and their corresponding definition in order are: Cg, copy gain; Rm, recurrent mutation; Nm, nominated mutation; Cm, clustered mutation; Oe, overexpression; and Hf, gene fusion.

To identify high level amplifications, the ‘*Cg*’ bit was set to 1 when the CN segment for a gene was estimated to exceed 6 total copies (log2 CN ≥1.5), the full gene was contained within the amplification, and the gene was detectably expressed (TPM ≥1). The expression filter prevents non-expressed genes within an amplified segment from being nominated as a potential oncogene.

To flag NS SNV/INDEL that likely result in a GOF events, we identified events that fell into one of three categories based on the altered amino acid position. To identify recurrent mutations, the ‘Rm’ bit is set for a gene in any patient where the observed mutation altered the same amino acid in at least two independent patients. When a gene was flagged as having recurrent mutations in the cohort, we also flagged clustered mutations in the gene to set the ‘*Cm*’ bit. Mutations were considered to be clustered when at least 5 different patients had NS mutations within 10% of the CDS space of a gene. Finally, when a gene was flagged as having recurrent mutations, we nominated any NS mutation in a gene by setting the ‘*Nm*’ bit (even including those without the ‘*Rm*’ or ‘*Cm*’ bits set) to capture potential GOF events by rare activating mutations.

To detect overexpression of a gene, which is common in myeloma through events like immunoglobulin translocations, we identified outlier RNA expression using the same MAD approach described in the LOF analysis. A gene was defined as overexpressed if the MAD value was greater than the 75th percentile value plus 1.5 times the interquartile range (IQR) and the observed corrected TPM ≥1. To limit false positive overexpression calls, we only set the ‘*Oe*’ bit when an accompanying DNA structural event was detected in the gene or its upstream or downstream region.

When an inframe fusion transcript was detected in the RNAseq data we set the ‘*Hf*’ bit for both genes involved in the fusion. The fusion events included were independently validated in the matching WGS assay, and the expression of both genes needed to be ≥1 TPM.

A gene was defined as gained in a sample if any of the ‘*Cg*’, ‘*Rm*’, ‘*Cm*’, ‘*Oe*’, or ‘*Hf*’ bits were set to 1. Samples with only the ‘*Nm*’ bit set for a gene were considered to have a potential GOF event and were only included in the count of samples with GOF events in a particular gene when at least 5 unique samples had one of the other bits set. In addition to the individual mechanism of gain, we also required the gene to be expressed (≥1 TPM) in the sample for it to be considered a GOF gene.

### Curation of LOF and GOF Genes

The integrated analysis focused on identifying genes with potential recurrent LOF and GOF events, however, some genes were nominated in both the GOF and LOF analysis (e.g. NRAS and KRAS). For genes that were identified as the target of both LOF and GOF in 5 or more patients, we relied on COSMIC Cancer Gene Census annotations (accessed August 2017) and reports of gene function from published literature. Known oncogenes removed from the LOF but retained on the GOF list were NRAS, KRAS, and WHSC1, while known tumor suppressor genes removed from the GOF list but retained on the LOF list were CDKN1B, DIS3, EGR1, FAM46C, MAX, NOTCH2, PABPC1, PARP4, RPL10, SDHA, TP53, and TRAF3. Genes that were exclusively nominated to the GOF list by mutational events were further curated based on their reported function in the literature and were removed if there was strong evidence supporting that the gene functions as a tumor suppressor rather than an oncogene. The genes removed through this process were DUSP2 and KLHL6, based on evidence from the literature, and BTG1, which is listed in the Cancer Gene Consensus as a tumor suppressor gene.

In patients where a gene had complete LOF as a result of two mutations, visual validation of mutations within close proximity was performed to determine if the mutations were in cis or trans. In order to perform visual validation, we required that sequencing reads or read-pairs spanned both of the mutation positions. If the mutations occurred in trans, no action was taken, however, if the mutations occurred in cis the gene was downgraded from complete to partial LOF for that particular sample. CDKN1B, EGR1, LRP6, MAGEC3, PABPC1, PRR14L, SYNE1, and UBR5 had samples whose variants were phased in cis, and after downgrading these events from complete to partial LOF, were removed from the list of recurrent complete LOF events occurring in 5 or more patients.

To further curate the LOF list, visual validation was performed to remove double calling events that can arise such as when a copy number deletion breakend overlaps a translocation breakpoint. Visual verification was limited to samples with contiguous genes identified as having a LOF involving a structural event. From the LOF list, CDKN2C, PSPC1, RB1, RCBTB2, CDC42BPB, CYLD, SNX20 had samples with LOF events that were manually modified after visual inspection. In addition, MAGI1 was removed from the GOF list because four of the five patients were flagged due to a translocation event in SLC25A26 which is believed to be a false-positive arising due to poor read mapping in this area.

### Identification of Copy Number Subtypes in Multiple Myeloma

CN subtype discovery and membership of the BM baseline samples was determined using the Monte Carlo reference-based consensus clustering tool M3C (v3.9) using the PAM clustering algorithm, Euclidean distance, and a max K of 20. In total, WGS data from 871 BM baseline tumor-normal pairs met quality control requirements for use in CN analysis. A uniform data matrix was created by extracting the largest overlapping CN state value for 26,771 non-overlapping 100 kb intervals that excluded centromere, immunoglobulin, and HLA regions along with sex chromosomes to remove biological and sex bias. To confirm the accuracy of the clustering results we ran three replicates, each with 200 iterations of consensus clustering using M3C, with the seed changing between each run.

### Identification of RNA Subtypes in Multiple Myeloma

Prior to unsupervised mRNA expression clustering, the log2(TPM+1) transformed expression data of 56,430 genes and 714 BM baseline tumor samples were adjusted for confounding biobank effects by applying ComBat from the sva R package (v3.26.0)^34^. The original list of genes were then ranked by their corresponding geometric coefficient of variation (GCV).

Larger GCV values are associated with greater variability in random variables that follow a log-normal distribution and therefore genes with a GCV less than 1 were removed from the dataset. The resulting 4811 gene by 714 sample expression matrix was mean centered as a method for standardizing the data. Clustering analysis was performed by running three separate trials of ConsensusClusterPlus (v1.42.0) with the PAM algorithm and Pearson distance metric^35^. Each trial computed sample class assignments for an increasing number of clusters (K), from 2 to 20, using the average linkage option and 80% subsampling with the number of repetitions set to 10,000. The optimum K was chosen based on the silhouette score for each cluster across all trials. All three trials with different initial seed values (3000094, 2862787, 8275806) converged to the solution K* = 12 by silhouette score.

Specific gene markers for each group were then determined using multinomial logistic regression with L1 regularization trained to predict class assignment^36^. For training, the model took the log2(TPM+1) mean centered expression matrix of the 4811 genes and 714 samples as input and class assignments from the unsupervised clustering as targets. The value of the regularization parameter that minimized the model’s objective function was determined during training via 20 fold cross-validation. The resulting non-zero values for the model coefficients were indicative of genes that were most predictive for a given class; 607 genes in total (Table S7).

Further analysis of the resulting RNA clusters included cross-examination with: a gene expression profiling index, used to assess cellular proliferation; an NFkB index; and previously identified RNA subtypes^17, 18, 28, 37, 38^. The proliferation index developed by Bergsagel et al. was calculated for each sample by measuring the average log2(TPM+1) transformed expression of 12 genes: TYMS, TK1, CCNB1, MKI67, KIAA101, KIAA0186, CKS1B, TOP2A, UBE2C, ZWINT,

TRIP13, KIF11. In addition, the NFkB(11) index for each sample was determined by calculating the geometric mean expression of 11 genes: BIRC3, TNFAIP3, NFKB2, IL2RG, NFKB1, RELB, NFKBIA, CD74, PLEK, MALT1, and WNT10A. Each subtype specific expression signature is composed of a list of up-regulated and down-regulated genes. In both Broyl and Zhan et al., these genes were identified using Affymetrix U133Plus2.0 microarray data. In order to apply these gene lists to RNAseq data, we identified the corresponding Ensembl v74 ENSG ID for each Affymetrix probe set using bioMartR, however, not all probesets had unique mappings to ENSG IDs. Many probesets were associated with multiple ENSG while other probesets were not associated with an ENSG. Only genes with unique probeset to ENSG mappings were used to calculate subtype specific expression signatures. Once the unique mappings were identified, each subtype specific signature was calculated for each sample using: *mean*(*log*_2_(*x*_*up*−*regulated*_) − (*x*_*down*−*regulated*_)). Integration of the genes predictive of each CoMMpass subtype, canonical structural events in MM, common CN events, previous subtype signatures, and expression index data was used to name the 12 subtypes.

To assign longitudinal samples with RNAseq data to each of these classes we developed a trained model classifier, which has the benefit of outputting class probabilities that can be used to evaluate transitions in time. The trained classifier uses the corrected TPM values for the 4811 genes used for clustering as input. These values are transformed, log2(TPM+1), followed by mean centering using the pre-computed mean expression values from the training set. Running this input through the trained classifier resulted in classifications for each of the 107 samples as well as the corresponding class probabilities associated with each of the 12 classes.

To determine whether checkpoint inhibitor and immunotherapy targets were expressed in PR patients, we compared the expression of each target in 51 PR patients and 663 non-PR patients at baseline. Targets for which the median expression in PR and non-PR patients was <1 TPM were excluded. An unpaired, two-sided Wilcoxon (Mann-Whitney) test was used to test the null hypothesis of equal median expression in both groups.

### Clinical and Molecular Associations with RNA subtypes

We used the non-parametric Fisher’s Exact test to determine if a clinical feature, LOF, or GOF event was enriched in the identified CN and RNA subtypes. The test works on the null table (Table S1), and the gene model. The two-tailed test was used to check for enrichment or depletion of each measurement within each subtype. An odds ratio was computed that indicated significant enrichment or depletion of the measurement in each subtype such that an odds ratio >1 indicated enrichment and <1 indicated depletion. In addition, for a condition to be depleted in a group it was required to be present in at least 20% of the cohort. The Benjamini-Hochberg test correction was used for multiple testing, which computed a pFDR value.

## Data Availability

DNA and RNA sequencing data is available from dbGAP, phs000748, and the Genomic Data Commons. Summarized somatic and clinical data used in this study are also available on the MMRF researcher gateway (https://research.themmrf.org/). Clinical data is updated and new molecular data is added with each bi-annual interim release.

https://www.ncbi.nlm.nih.gov/projects/gap/cgi-bin/study.cgi?study_id=phs000748.v7.p4

https://portal.gdc.cancer.gov/

## Acknowledgements

The CoMMpass study was funded by the Multiple Myeloma Research Foundation. We would like to thank all of the MMRF CoMMpass study participants and their families for making this research possible. We would also like to thank all of the laboratory staff at the Spectrum Health Advanced Technology Laboratory who assisted with sample processing and flow cytometry analysis.

## Data Availability

DNA and RNA sequencing data are available from dbGAP, phs000748, and the Genomic Data Commons. Summarized somatic and clinical data are also available on the MMRF researcher gateway (https://research.themmrf.org/). Clinical data is updated and new molecular data is added with each bi-annual interim release.

## Author Contributions

Conceptualization, S.S, D.A., J.C., S.L., J.K.; Methodology, S.S., D.P., A.W.C., S.N., M.B., S.K., K.S., D.C., W.L., J.K.; Software, S.S., D.P., A.W.C., S.N., J.L.A., C.L., B.B., C.M., B.T., A.K., M.W., V.Y., M.T., K.M., D.C., J.K.; Validation, S.S., A.W.C., M.T., K.M., J.K.; Formal Analysis, S.S., D.P., A.W.C., S.N., J.L.A., C.L., B.B., C.M., B.T., A.K., M.W., V.Y., J.K.; Investigation, J.R.A., L.C., M.B., A.H., S.K., J.M., R.R., K.S., E.T., A.B., B.D., M.K., M.D.A., M.G., J.S., P.K., A.D., B.Z., M.T., K.M., H.J.C., A.C., S.J., D.S.S., R.V., G.O., A.J., R.N., D.L., M.L., J.B., S.Z.U., D.C., S.D.J., S.L., J.K.; Resources, H.J.C., S.J., D.S.S., R.V., G.O., A.J., R.N., D.L., M.L., J.B., S.L.; Data Curation, S.S., D.P., A.W.C., S.N., J.L.A., C.L., D.C.R., M.D.A., M.G., A.D., B.Z., M.T., K.M., M.D., H.J.C., J.Y., D.A., S.D.J., J.K.; Writing - Original Draft, S.S., J.K.; Writing - Review & Editing S.S., B.B., D.S.S., N.C.G., A.C., S.Z.U., K.C.A., H.J.C., J.K.; Visualization, S.S., D.P., A.W.C., S.N., J.L.A.; Supervision, S.S., D.C., J.Y., D.A., S.D.J., J.C., S.L., J.K.; Project Administration, D.C.R., D.C., A.D., B.Z., K.M., M.D., J.Y., D.A., N.C.G., S.D.J., J.C., S.L., J.K.; Funding Acquisition, P.K., S.D.J., J.C., J.K.

