## Supplemental Figures for "Genomic Basis of Multiple Myeloma Subtypes from the MMRF CoMMpass Study"

#### Table of Contents

**1 Cohort Description** 2
1.1 Survival Outcomes of the Cohort 2
1.2 Survival Outcomes of Patients with High Risk Molecular Features 3
1.3 Baseline Molecular Assay Intersection 4

**2 Identification of DNA Subtypes of Multiple Myeloma** 5
2.1 Cluster Number Determination from CN Consensus Clustering 5
2.2 Consistency of Subtype Assignments by CN Clustering 6
2.3 CN Consensus Clustering Matrix 7
2.4 CN Density Plots for Subtype Defining Events 8
2.5 Survival Outcomes for HRD and NHRD Patients 9
2.6 Survival Outcomes for Patients with Amp1q and Del13q 10

**3 Identification of RNA Subtypes of Multiple Myeloma** 11
3.1 Cluster Number Determination from Gene Expression Consensus Clustering 11
3.2 Consistency of Subtype Assignments by RNA Expression Clustering 12
3.3 RNAseq Consensus Clustering Matrix 13
3.4 RNA Subtypes and Association with Copy Number 14
3.5 Relationship between CoMMpass and Zhan et al. Expression Subtypes 15
3.6 Relationship between CoMMpass and Broyl et al. Expression Subtypes 16
3.7 CCND1/2/3 and PAX5 Expression Across RNA Subtypes 17
3.8 Relationship between Proliferation Index and CoMMpass Subtypes 18
3.9 Prevalence of Bone Disease Across RNA Subtypes 19
3.10 NFKB Index Distribution by RNA Subtype 20
3.11 NINJ1 and TP53 Expression Across RNA Subtypes 21
3.12 Low Purity Association with Low Purity Metrics 22

**4 Clinical and Molecular Associations with RNA Subtypes** 23
4.1 Mechanisms of RB1 Complete Loss at Diagnosis 23

**5 Transition to PR at Progression and Link with G1/S** 24
5.1 Change in RNA Subtype Probabilities Over Time 24
5.2 Overall Survival of Patients After Transition to PR Subtype 25
5.3 Deletion of of CDKN2C in Patients that Transitioned to PR 26
5.4 Deletion of of CDKN1B in Patient that Transitioned to PR 27

### 1 Cohort Description

#### 1.1 Survival Outcomes of the Cohort

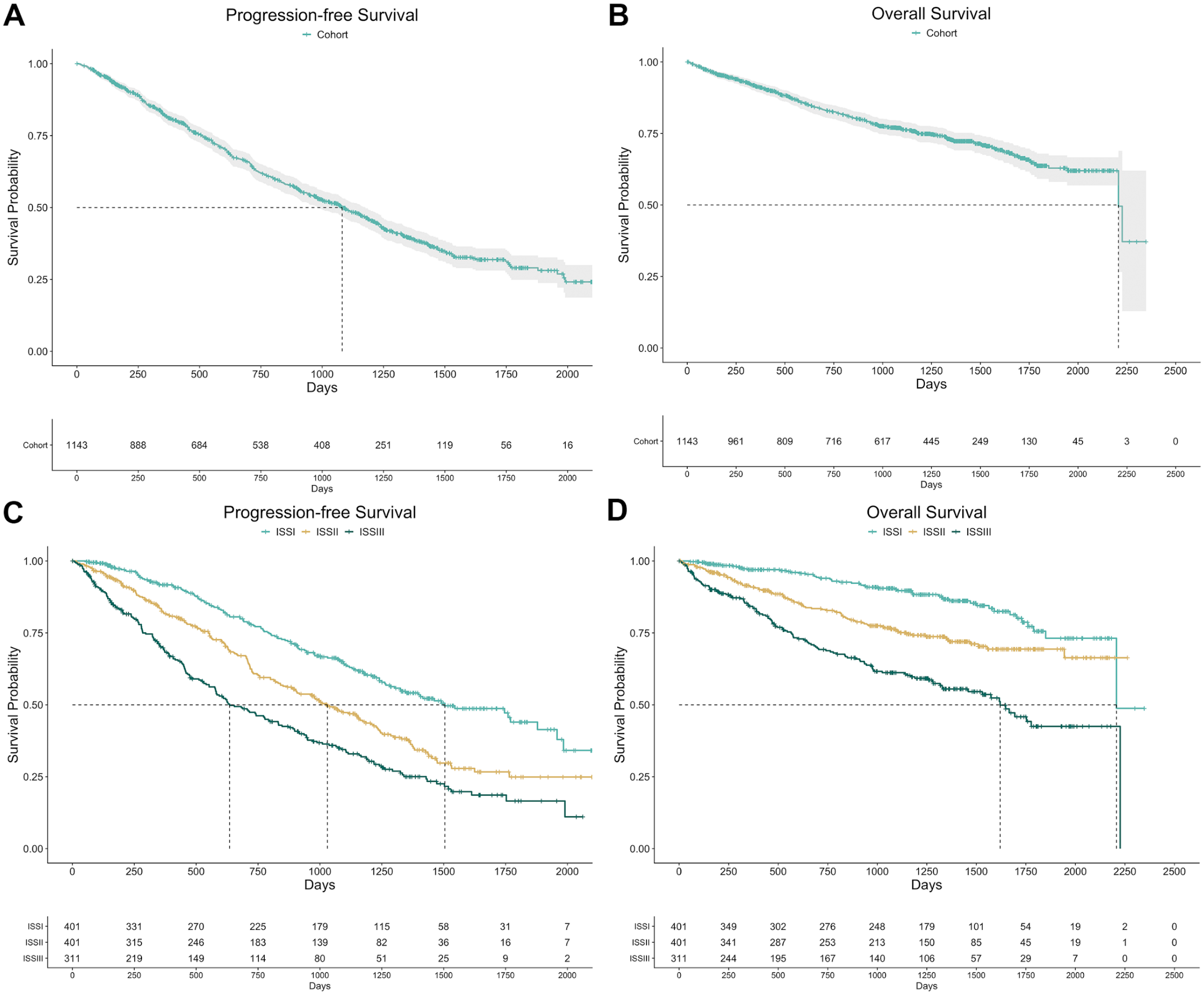


Progression-free (PFS) and overall (OS) survival outcomes of the CoMMpass cohort (A-B) and the CoMMpass cohort stratified by ISS stage (C-D). (A-B) Median PFS (36 months) and OS (74 months) of the cohort has been met, however as of the IA14 release there is insufficient cohort follow up to accurately report median cohort OS within a 95% confidence interval. (C) PFS outcomes for patients classified as ISSI (50 months), ISSII (34 months), and ISSIII (21 months) at diagnosis. (D) OS outcomes for patients classified as ISSI (74 months), ISSII (median not met), and ISSIII (54 months) at diagnosis. ISS stage stratified patients into three clinically distinct classes, with patients classified as ISSII at diagnosis having poor OS and PFS outcomes as compared to patients classified as ISSI ($p < 0.001$), and patients classified as ISSIII having poor PFS outcomes as compared to patients classified as ISSII ($p < 0.001$).

#### 1.2 Survival Outcomes of Patients with High Risk Molecular Features

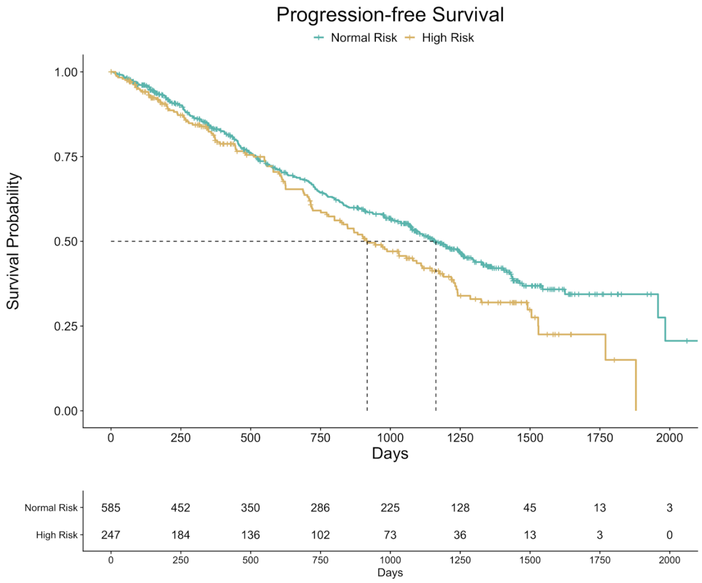


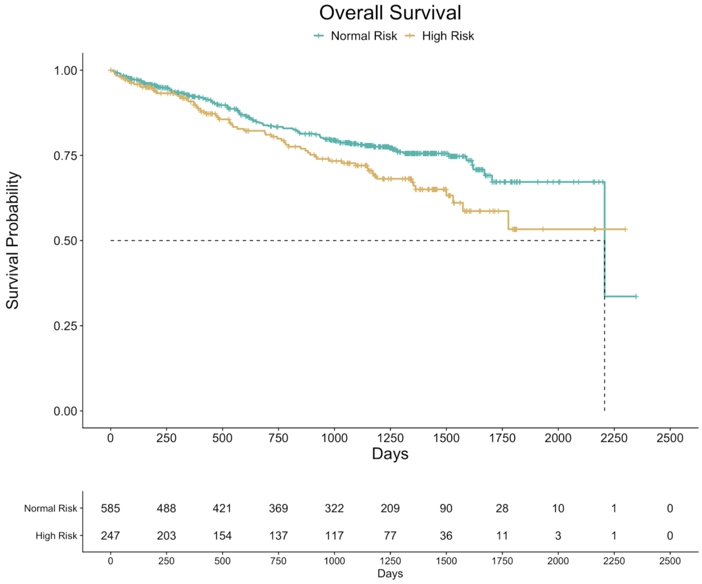


PFS and OS outcomes of CoMMpass patients with high risk molecular features: del17p13, t(14;16) MAF, t(14;20) MAFB, t(8;14) MAFA, or t(4;14) WHSC1/MMSET/NSD2. Median PFS for normal-risk and high-risk patients was 39 and 31 months respectively. Median OS for normal-risk patients was 73.6 months, whereas median OS for high-risk patients was not met as of the IA14 release. Patients with high-risk features had poor PFS and OS outcomes ($p < 0.05$) as compared to normal-risk patients.

#### 1.3 Baseline Molecular Assay Intersection

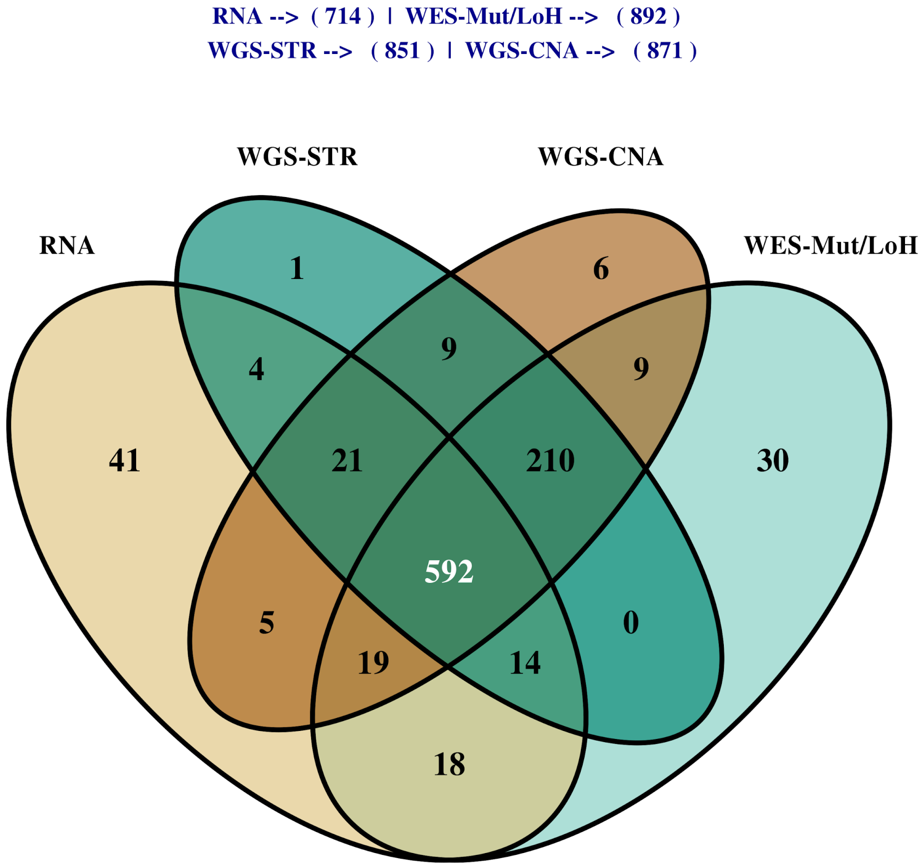


Venn diagram showing the number of patients in the baseline CoMMpass cohort with available RNAseq, WGS structural (WGS-STR), WGS copy number (WGS-CNA), and WES mutation and loss-of-heterozygosity (WES-Mut/LoH) data for bone marrow (BM) derived tumor samples. There were 592 BM-derived tumor samples that were fully characterized with all data types available for analysis at diagnosis.

### 2 Identification of DNA Subtypes of Multiple Myeloma

#### 2.1 Cluster Number Determination from CN Consensus Clustering

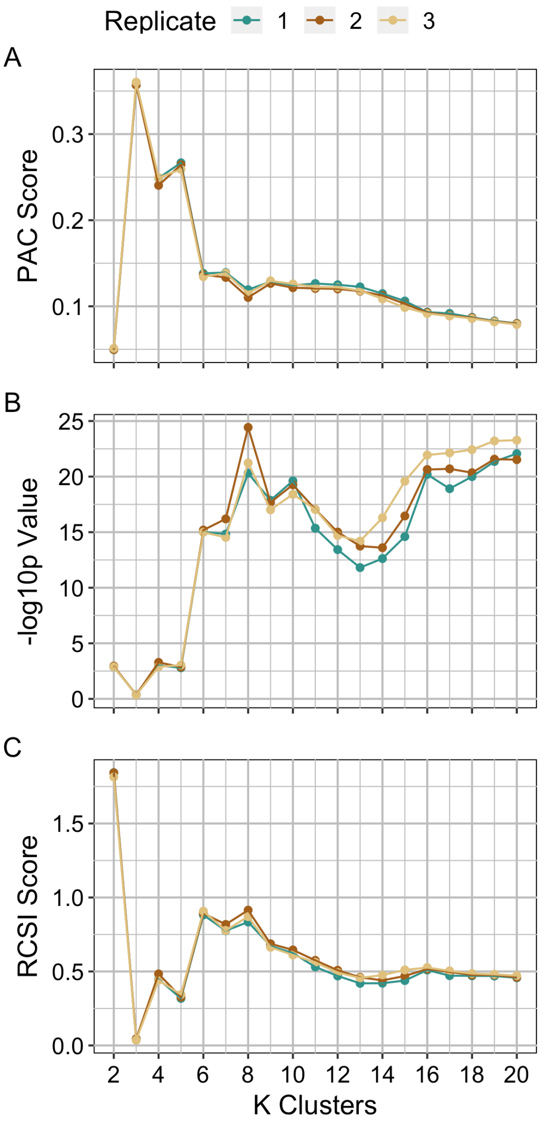


Consensus clustering was performed in triplicate using three different seeds and the optimum number of clusters was determined using M3C. M3C calculates (A) a PAC score, (B) -log10p value, and (C) RCSI score for K=2 to maxK. A lower PAC score, higher -log10p value, and higher RCSI score indicate an optimal number of clusters. An optimal class number of K=2 is supported by both the PAC score and the RCSI, with the lowest PAC score and the highest RCSI score across all three replicates, identifying the two broad HRD and NHRD genetic subtypes of myeloma. However, the p-value for K=2 is among the lowest when compared to the other clusters. For values of K greater than 12, the PAC score and p-score begin to overfit the data, indicated by the downward and upward trend respectively. Within the range of K=3-12, K=8 has the lowest PAC score, most significant p value, and a consistently high RCSI score across all three replicates, indicating K=8 is the optimal cluster number. The class assignments from replicate 2 were used for all further downstream analyses.

#### 2.2 Consistency of Subtype Assignments by CN Clustering

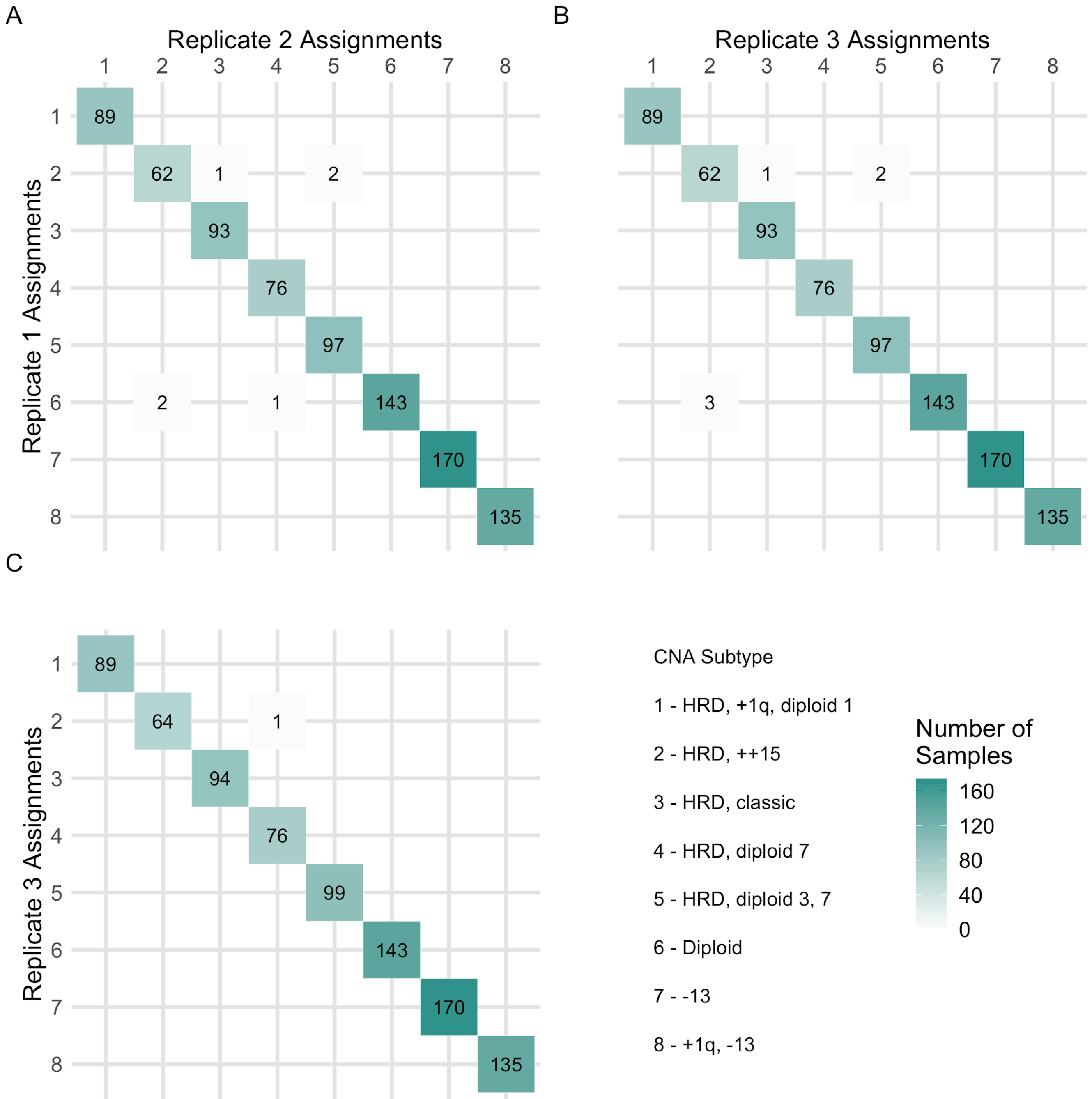


Confusion matrices showing the number of patients classified in each CN subtype across three replicates. CN subtype classifications were highly consistent across replicates with only 6/871 (0.7%) patients having different CN subtype classifications across replicates (A) 1 and 2 and (B) 1 and 3, and (C) only 1/871 (0.1%) patients having a different CN subtype classification across replicate 2 and 3.

#### 2.3 CN Consensus Clustering Matrix

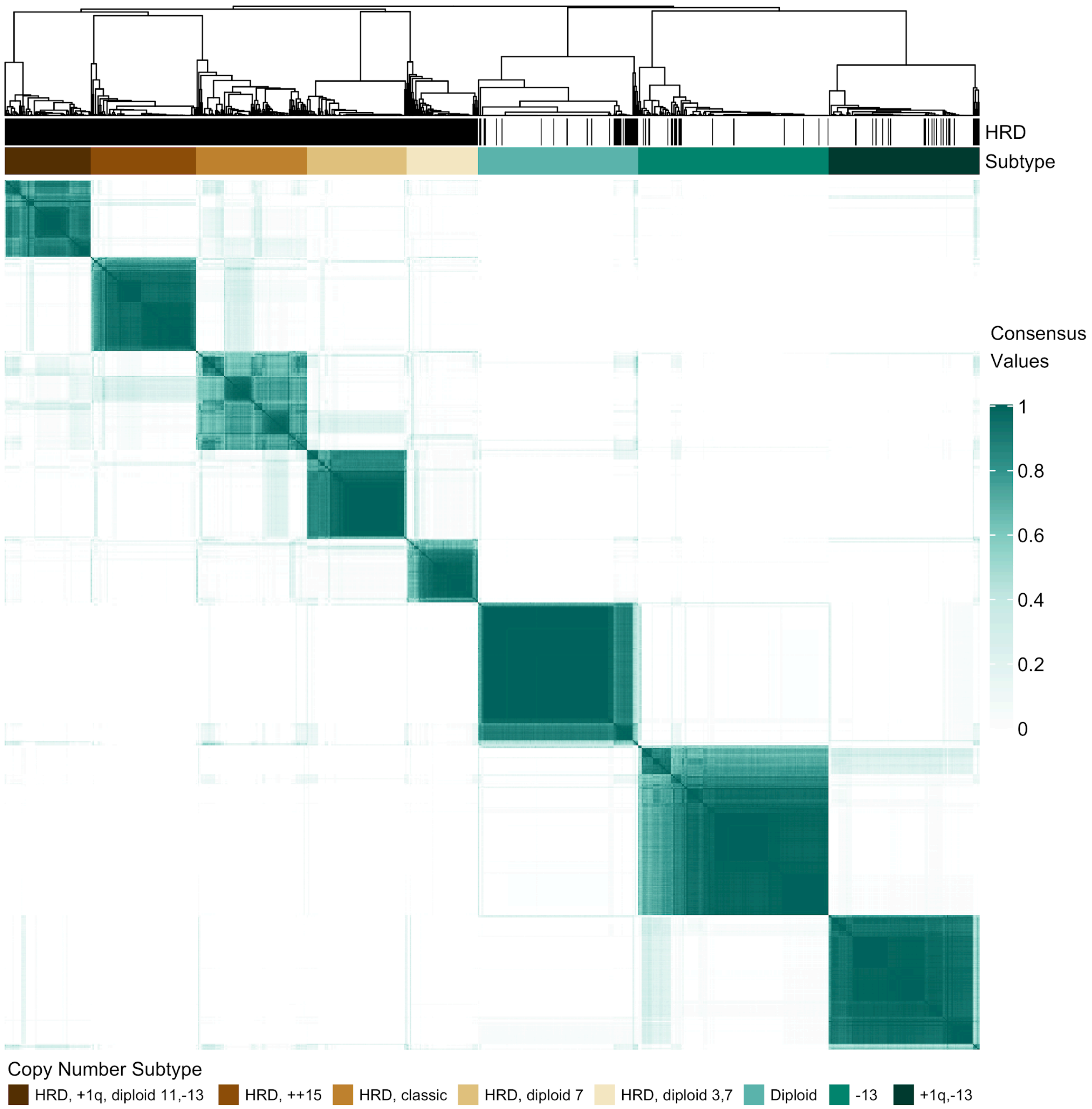


Consensus clustering matrix with an optimal clustering solution of K=8. The M3C (Monte Carlo reference-based) consensus clustering algorithm was applied to the CN measurements of 26,771 100Kb intervals across the GRCh37 reference genome for 871 WGS BM derived baseline samples. Five of the eight subtypes include only samples classified as hyperdiploid.

#### 2.4 CN Density Plots for Subtype Defining Events

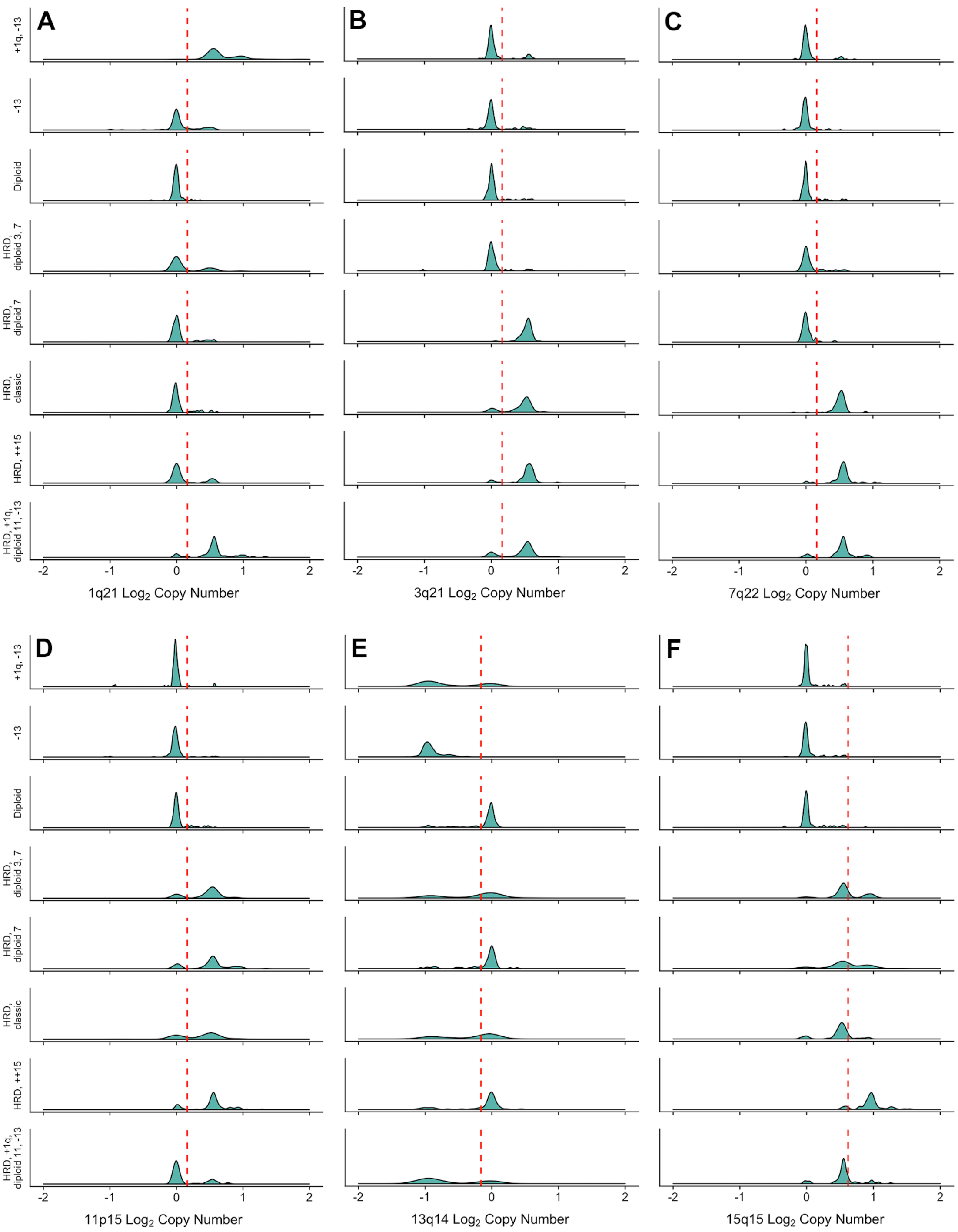


Copy number (log2) density plot across CN subtypes for (A) 1q21, (B) 3q21, (C) 7q22, (D) 11p15, (E) 13q14, and (F) 15q15. The red dotted line in plots A-D indicates the 1 copy gain threshold, in plot E indicates the 1 copy loss threshold, and in plot F indicates the 2 copy gain threshold.

#### 2.5 Survival Outcomes for HRD and NHRD Patients

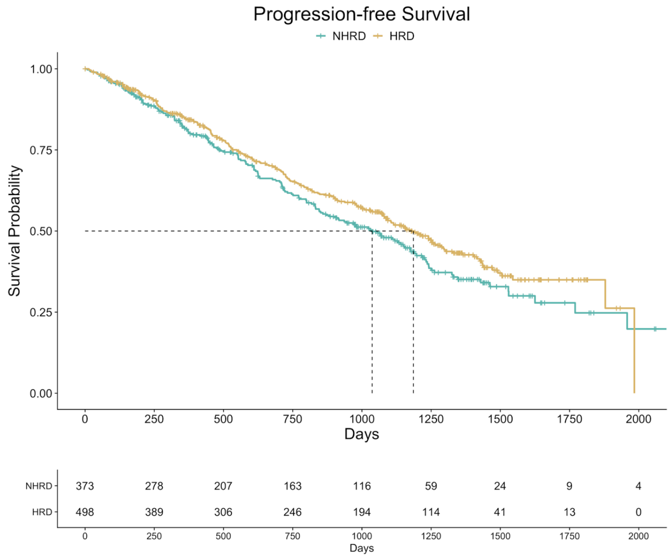


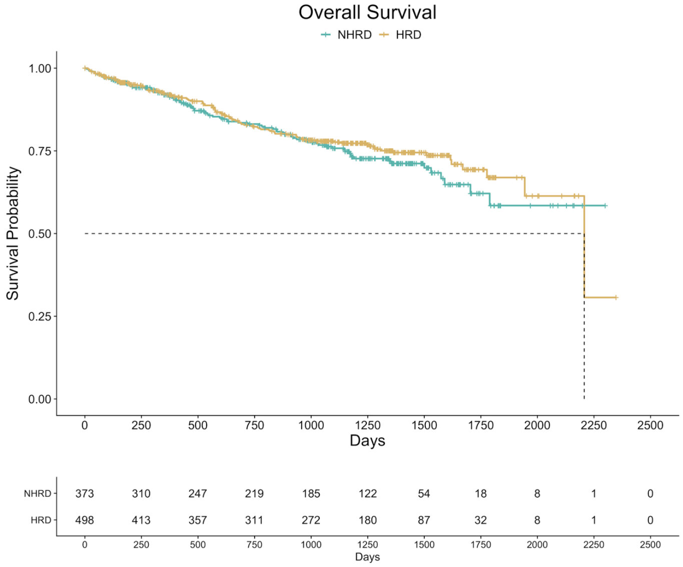


PFS and OS outcomes for hyperdiploid (HRD) versus non-hyperdiploid (NHRD) CoMMpass patients. There was no significant difference in PFS for HRD (39.5 months) versus NHRD (34.6 months) patients, or OS for HRD (73.6 months) versus NHRD (median OS not met) patients.

#### 2.6 Survival Outcomes for Patients with Amp1q and Del13q

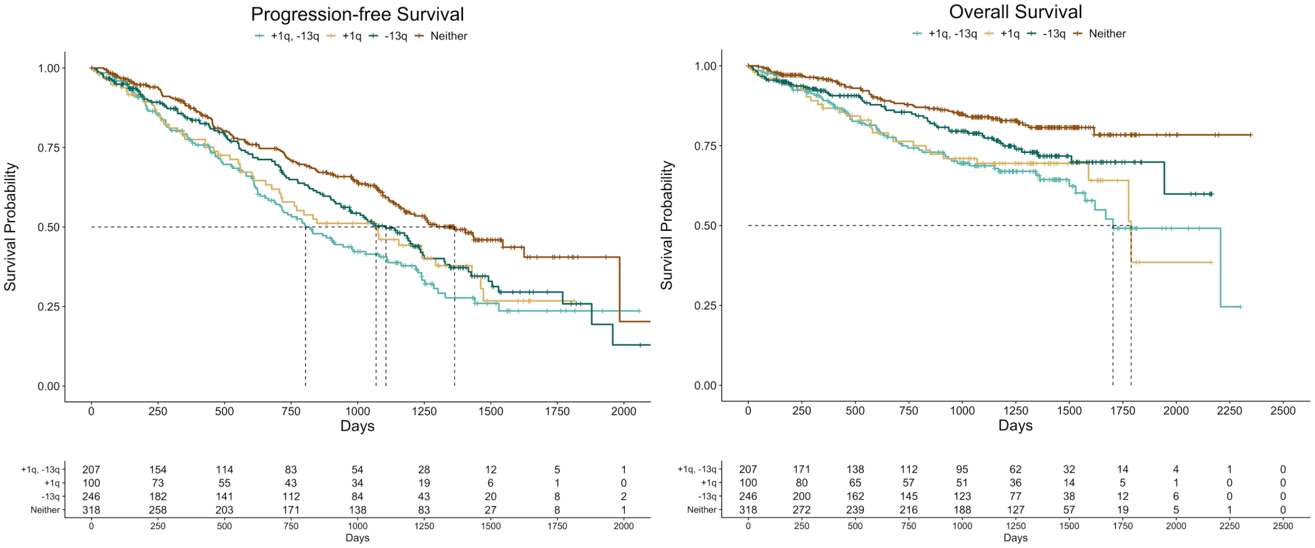


Univariate Cox Proportional Hazards Model

| Variable | No. Patients (%) | HR | 95% CI | p-value |
|---|---|---|---|---|
| Amp1q | 307 (35.2) | 1.8 | 1.4-2.4 | <0.0001 |
| Del13q | 453 (52.0) | 1.5 | 1.1-2.0 | 0.0044 |

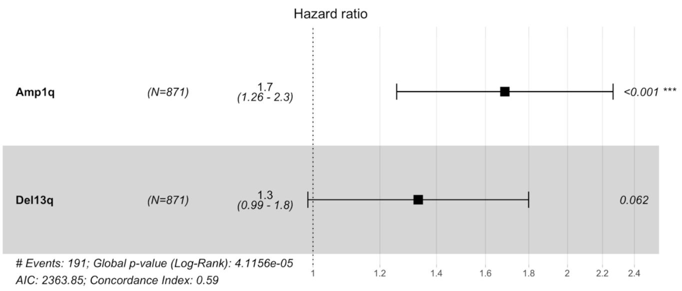


PFS (top left) and OS (top right) outcomes for CoMMpass patients with both amp1q and del13q (+1q21, -13q14), amp1q alone (+1q21), del13q alone (-13q14), and neither amp1q or del13q (neither). Amp1q was defined as a gain of 1 or more copies of 1q21, whereas del13q was defined as a loss of one copy of 13q14. No difference in PFS or OS outcome was observed between +1q21, -13q14 patients and +1q21 patients, however patients with +1q21, -13q14 had poor OS outcomes compared to patients with -13q14 alone ($p < 0.05$). In a univariate Cox proportional hazards model, both amp1q and del13q were found to significantly impact OS outcome (bottom left). However in a multivariate model (bottom right), only amp1q was found to have a significant impact on outcome after adjusting for del13q status.

### 3 Identification of RNA Subtypes of Multiple Myeloma

#### 3.1 Cluster Number Determination from Gene Expression Consensus Clustering

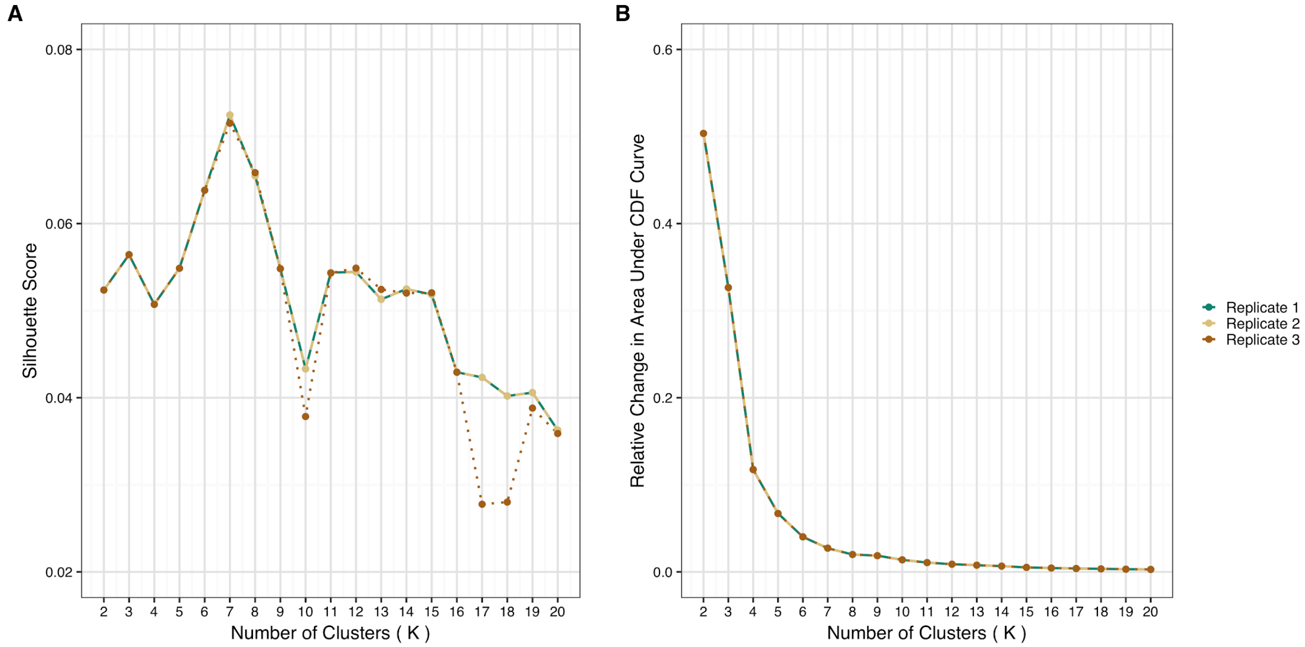


For each of the three replicates of consensus clustering, a (A) silhouette score and (B) relative change in area under the CDF curve was computed for the number of possible clusters tested (K, 2-20). (A) The silhouette score is defined as the average silhouette width, s(x), of all samples in the dataset, where s(x) is a measure of how appropriately grouped all samples are within a cluster, and the average of all silhouette widths reflects overall clustering quality. (B) The proportion increase in the CDF area as K increases, $\Delta K$. Ideally, the optimal number of clusters (K) will correspond to the K that maximizes the silhouette score, s(x), while minimizing the $\Delta K$. We evaluated the resulting grouping from K = 7-15 based on the aforementioned criteria (s(x) score and $\Delta K$) in combination with CoMMpass WGS data and groups identified in previous studies to identify biologically relevant subtypes. Previous studies identified four common groups: MS, CD1, CD2, and PR.  In our consensus clustering trials, these classes were not identified until K=11 or greater, in particular, the PR subtype which is the only group with a significant difference in outcome. We ultimately selected K=12, as it had the highest s(x) while minimizing $\Delta K$ when compared to other local maximums of s(x). K=9 was eliminated because one cluster only contained 2 patients, and similarly K=10 contained a cluster with only 1 patient, resulting in spurious groups that were not informative of myeloma biology.

#### 3.2 Consistency of Subtype Assignments by RNA Expression Clustering

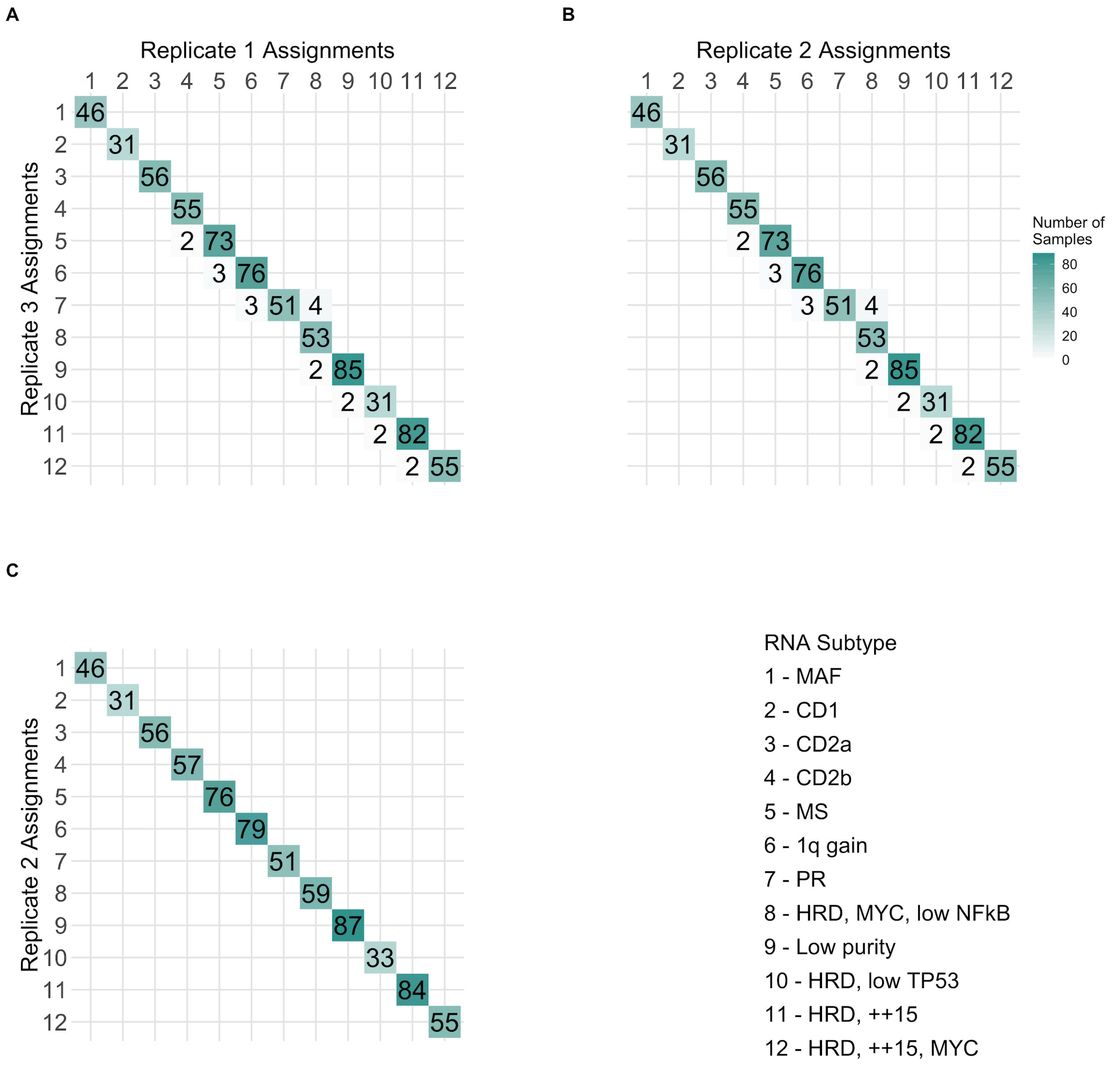


Confusion matrices of the number of patients classified in each RNA subtype across three replicates. (C) Replicate 1 and 2 produced identical cluster assignments for all 714 samples. (A-B) 20 samples (2.8% of the total dataset) from replicate 3 were discordant from their assignments in replicates 1 and 2.

#### 3.3 RNAseq Consensus Clustering Matrix

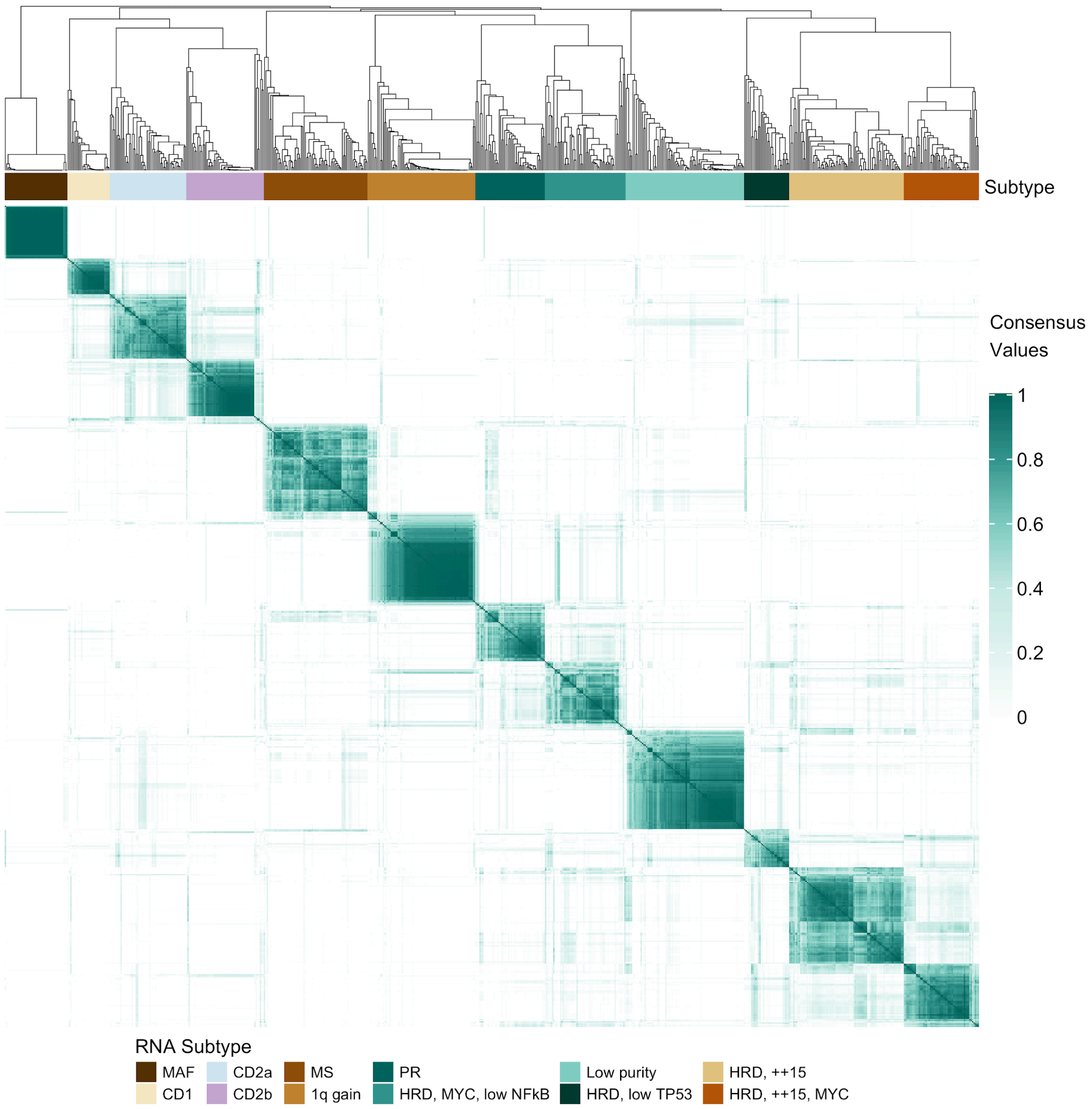


Consensus matrix showing the consistency of class assignment for K=12 clustering of RNA-seq data derived from 714 BM baseline samples and 4811 feature-selected genes.

#### 3.4 RNA Subtypes and Association with Copy Number

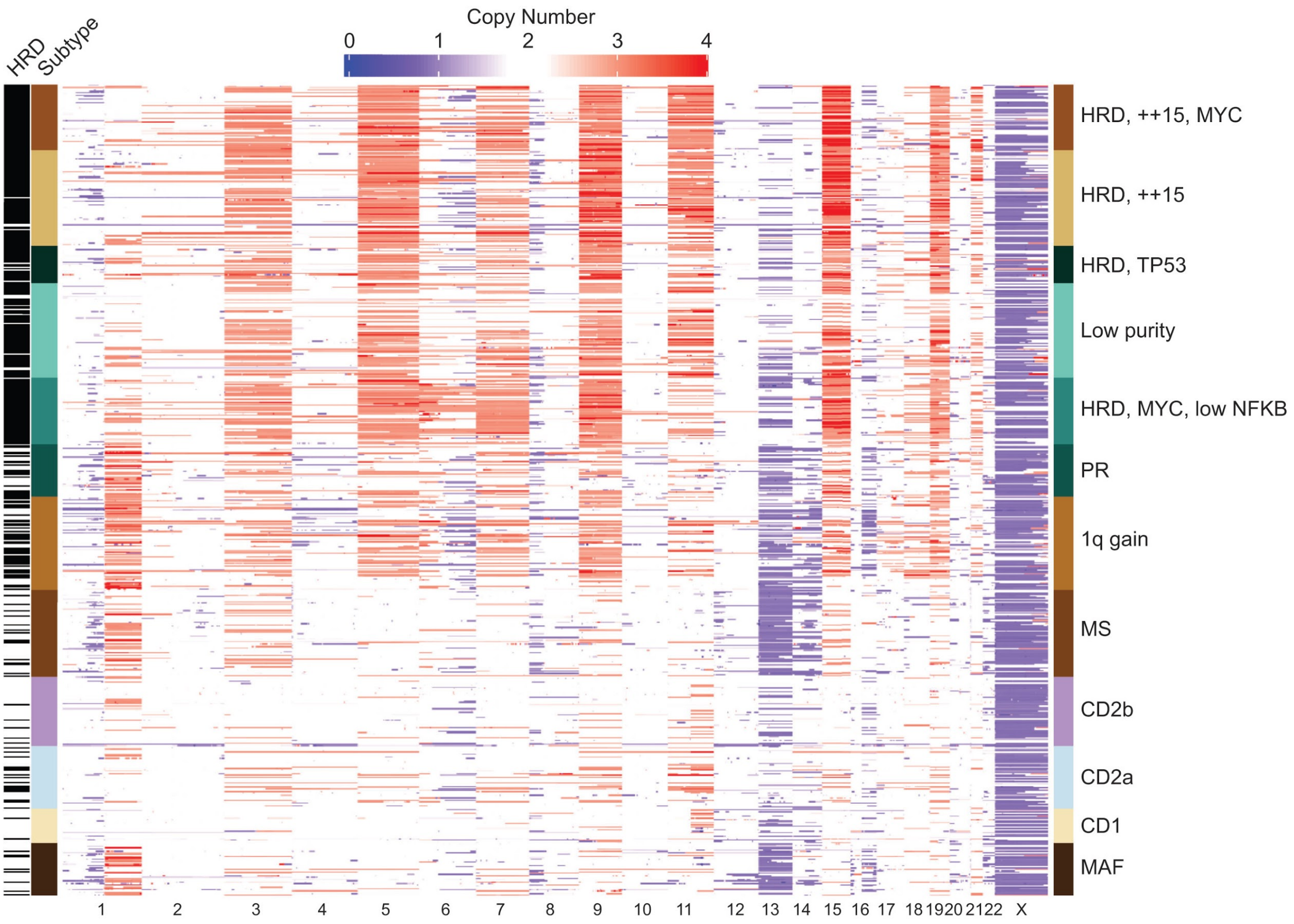


Copy number states for patients by RNA subtype are shown. Diploid copy number is represented as 2 (white), copy loss is shaded blue, and copy gain is shaded in red. Rare copy number values exceeding 4 are represented as a copy number value of 4 to maintain uniformity in the heatmap scales for gain and loss.

#### 3.5 Relationship between CoMMpass and Zhan et al. Expression Subtypes

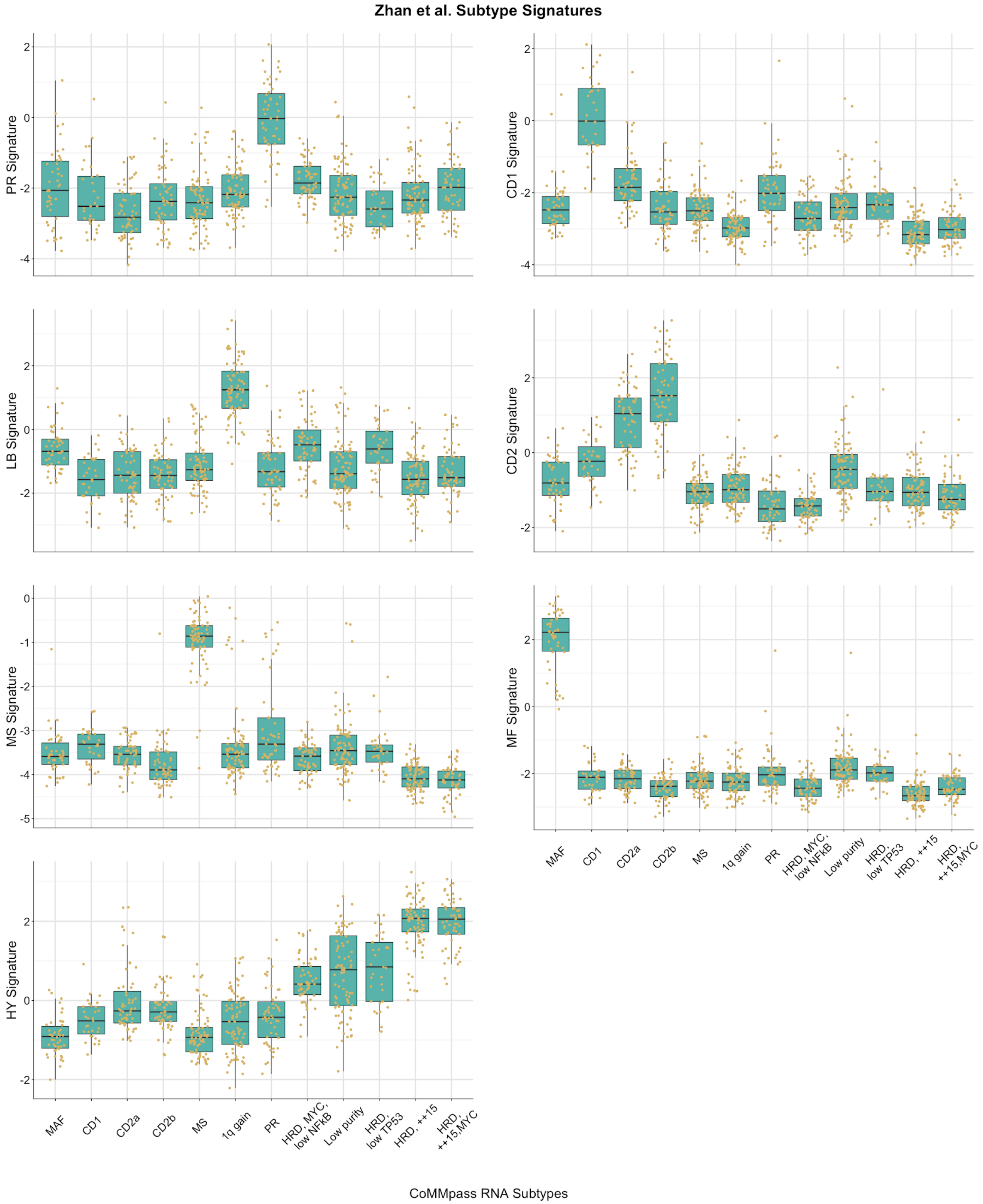


Index values for each of the 7 subtypes defined by Zhan et al.[17] were calculated for each patient and compared to the CoMMpass RNAseq native subtype assignments. The distribution of index values between the Zhan et al. subtypes and each identified CoMMpass subtype was used to identify related subtypes.

### 3.6 Relationship between CoMMpass and Broyl et al. Expression Subtypes

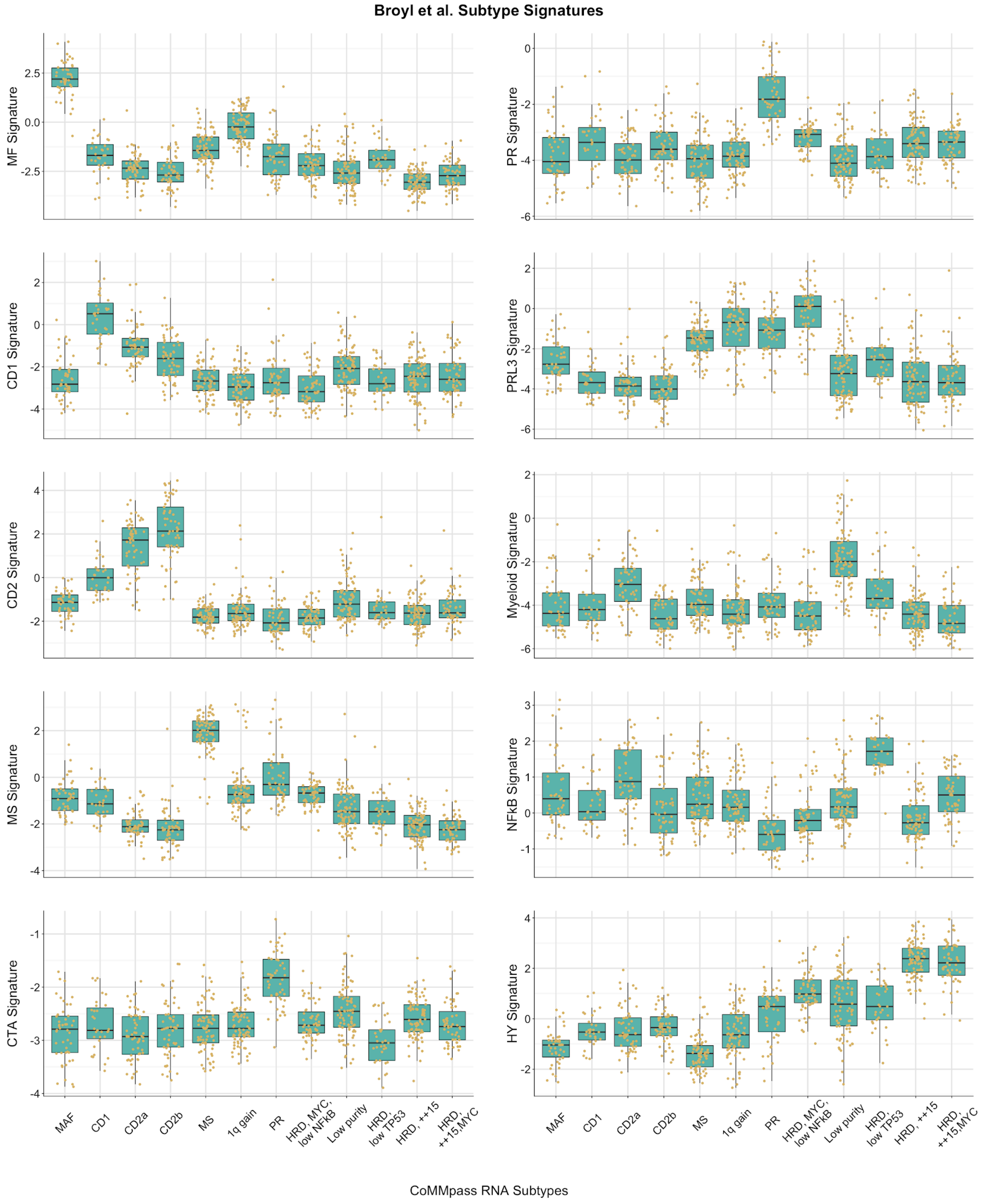


Index values for each of the 10 subtypes defined by Broyl et al.[18] were calculated for each patient and compared to the CoMMpass RNAseq native subtype assignments. The distribution of index values between the Broyl et al. subtypes and each identified CoMMpass subtype was used to identify related subtypes.

#### 3.7 CCND1/2/3 and PAX5 Expression Across RNA Subtypes

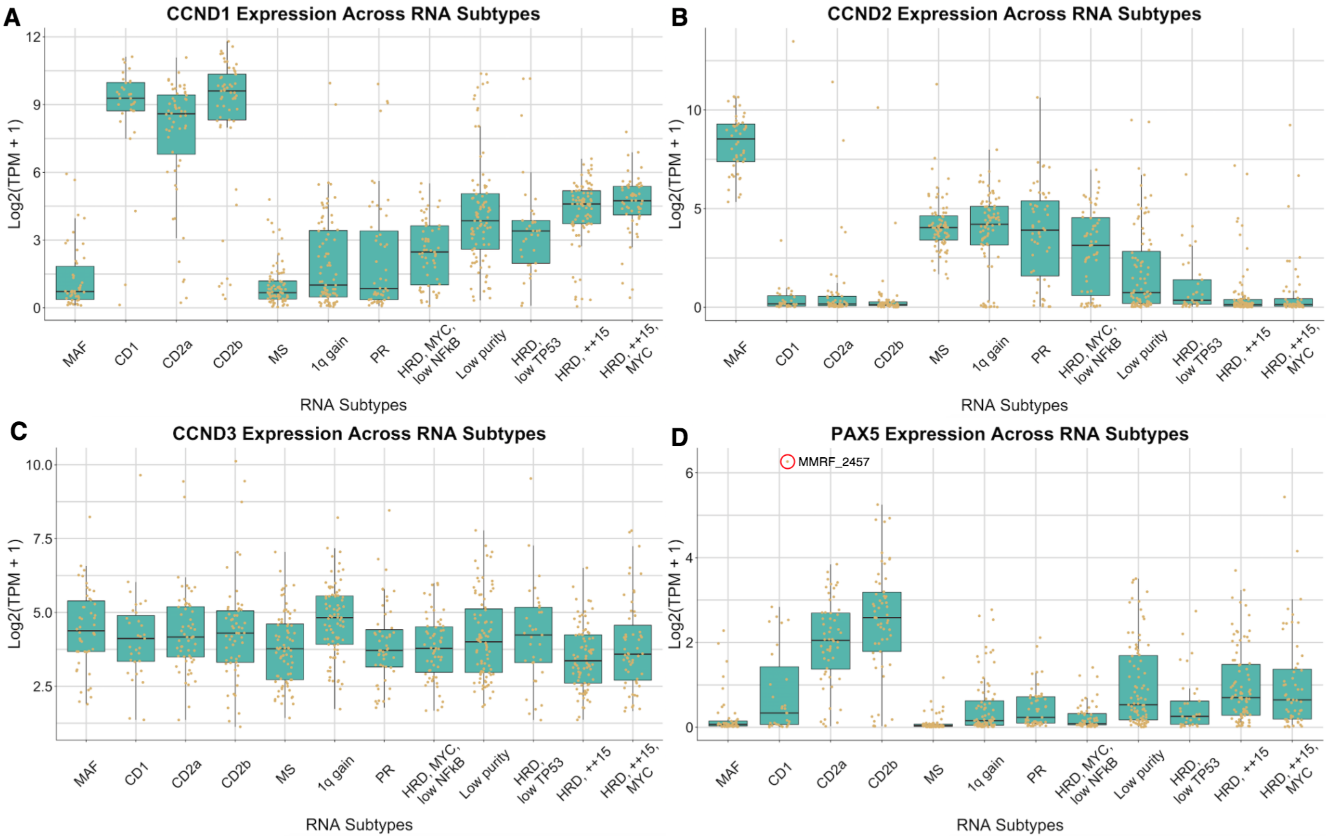


(A) CCND1, (B) CCND2, (C) CCND3, and (D) PAX5 expression across RNA subtypes. Patients (brown dots) in the CD1, CD2a, and CD2b subtypes typically had overexpression of CCND1, CCND2, or CCND3 due to canonical immunoglobulin transolocations targeting these genes. In MMRF_2457 (red circle), a translocation involving CCND1/2/3 was not identified, however, this patient had a t(9;14) resulting in overexpression of PAX5. Notably, the CD2a and CD2b subtypes had the highest median expression of PAX5 across RNA subtypes.

#### 3.8 Relationship between Proliferation Index and CoMMpass Subtypes

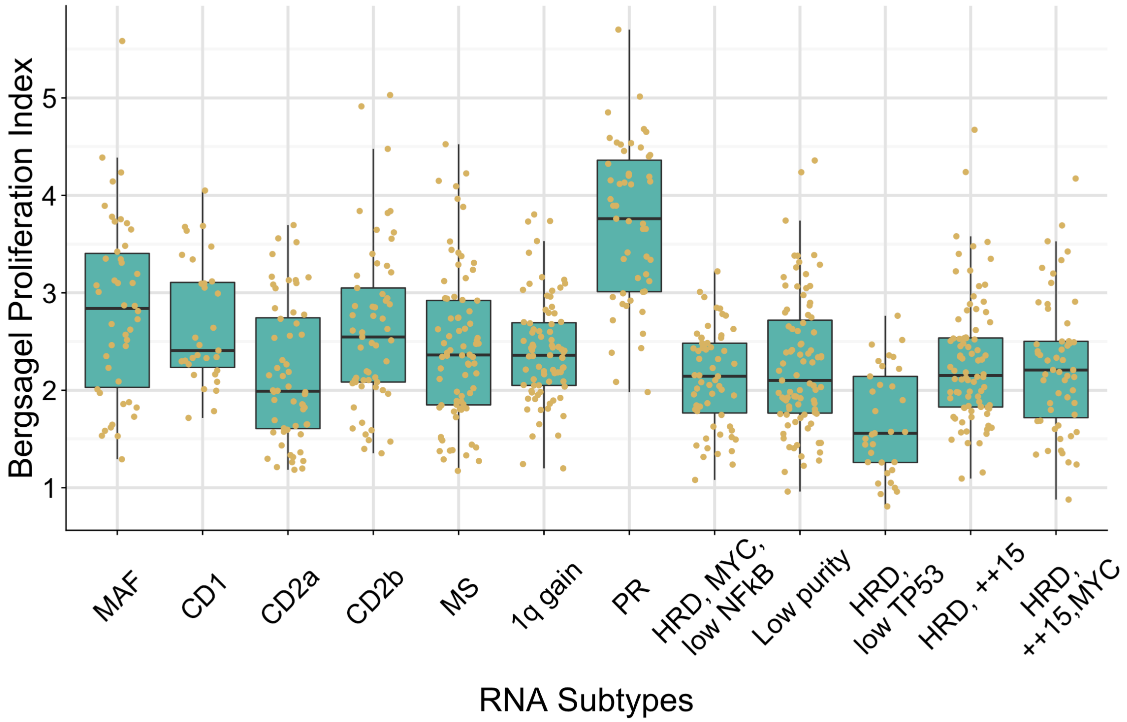


The association with an RNAseq defined proliferation index and CoMMpass subtypes is shown. The Bergsagel Proliferation Index[28] for each sample was determined by calculating the geometric mean expression of 12 genes (TYMS, TK1, CCNB1, MKI67, KIAA101, KIAA0186, CKS1B, TOP2A, UBE2C, ZWINT, TRIP13, KIF11). The PR subtype had the highest median proliferation index score.

#### 3.9 Prevalence of Bone Disease Across RNA Subtypes

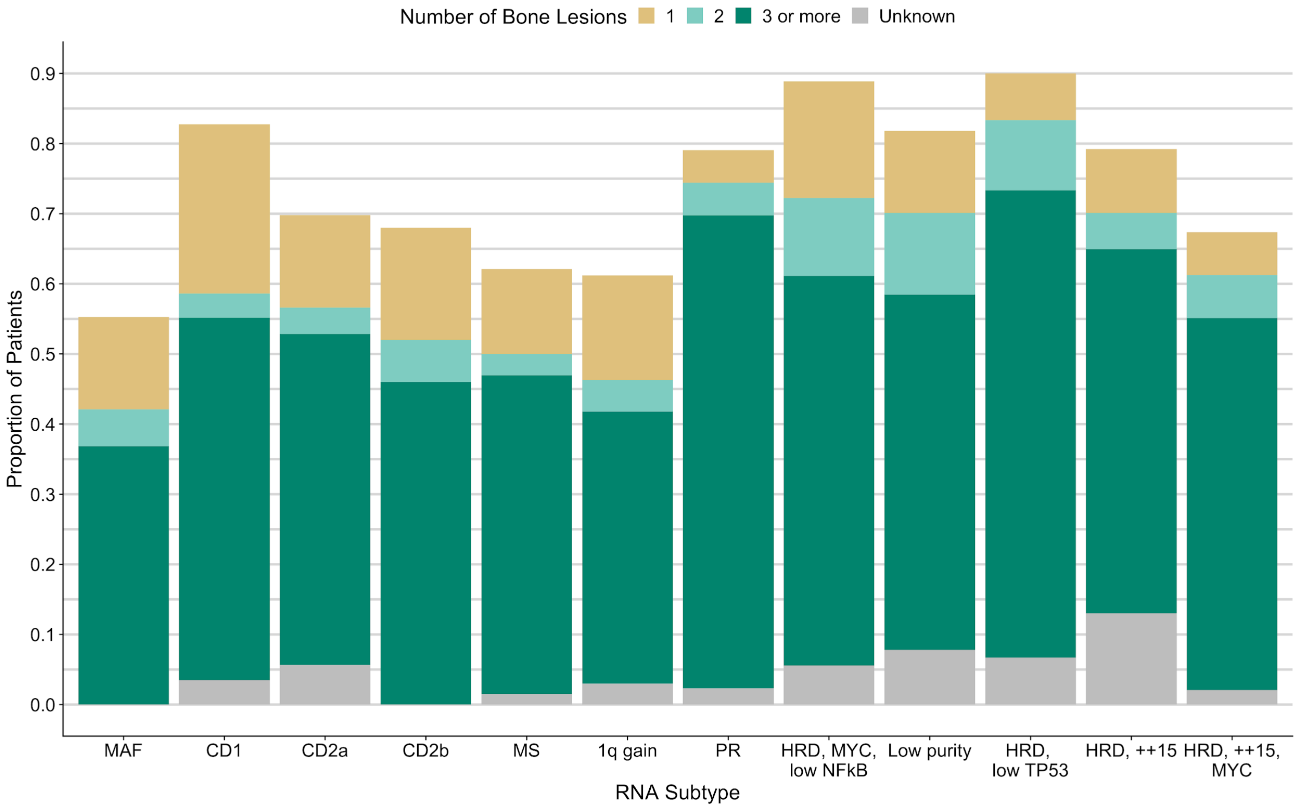


Proportion of patients in each RNA subtype with bone disease. The proportion of patients with 1, 2, or 3 or more bone lesions for each subtype is also shown. The Unknown (gray) category represents patients with bone disease for whom the number of lesions was not specified.

#### 3.10 NFKB Index Distribution by RNA Subtype

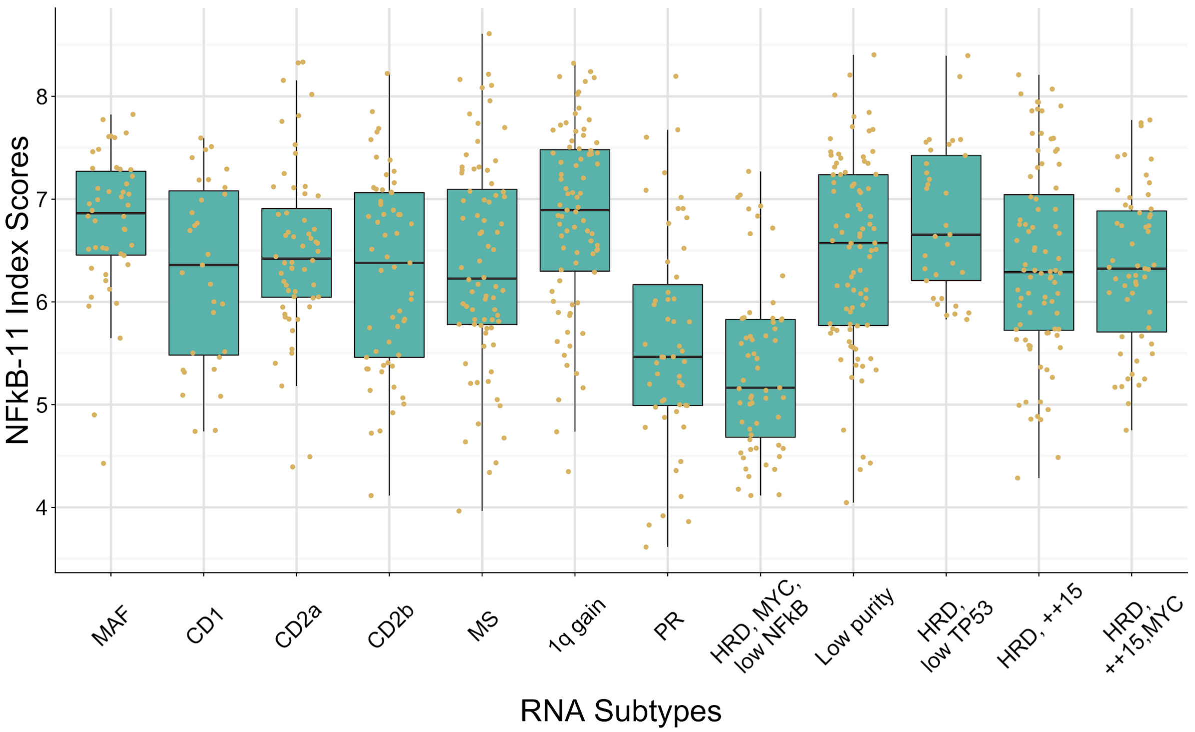


The association with an RNAseq defined NFKB index and the CoMMpass subtypes is shown. The NFKB(11) index for each sample was determined by calculating the geometric mean expression of 11 genes (BIRC3; TNFAIP3; NFKB2; IL2RG; NFKB1; RELB; NFKBIA; CD74; PLEK; MALT1; and WNT10A)[36,37].

#### 3.11 NINJ1 and TP53 Expression Across RNA Subtypes

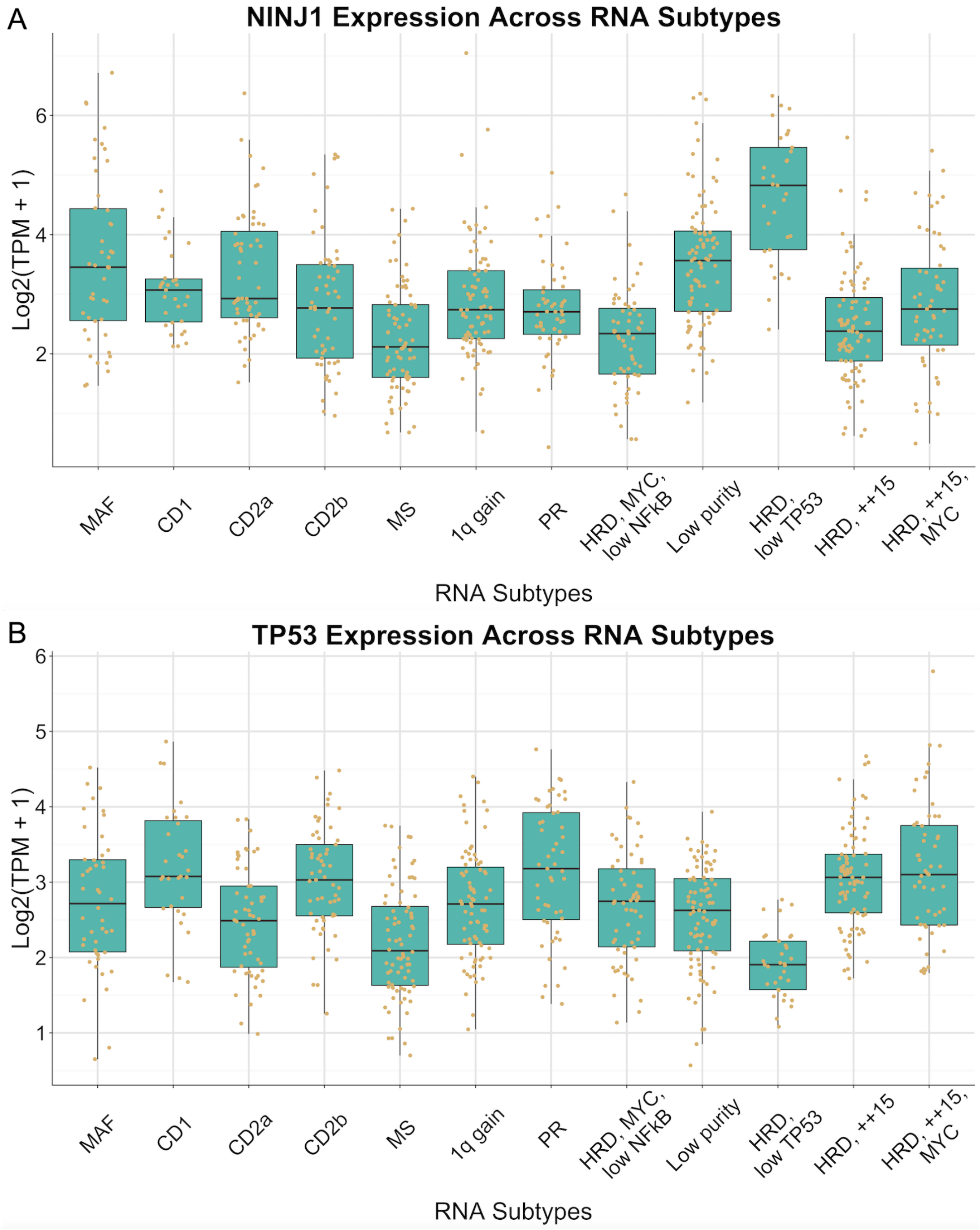


The expression of (A) NINJ1 and (B) TP53 is shown for each CoMMpass RNA subtype.

#### 3.12 Low Purity Association with Low Purity Metrics

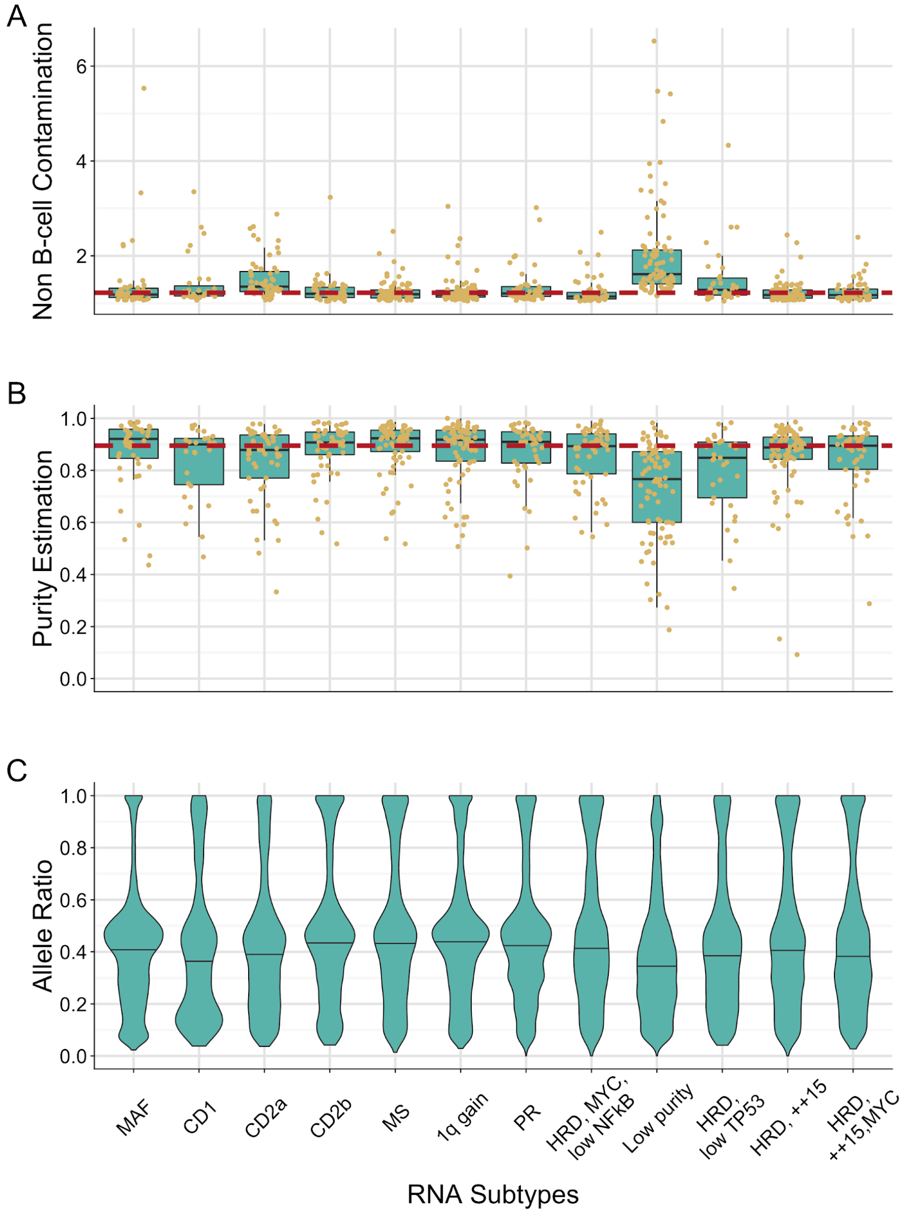


The low purity RNA subtype was defined based on an association of the samples in this category with multiple independent measures of sample purity. (A) An index associated with genes expressed in non B-cell tissues was used to identify samples with contamination of non-B lineage cells in the $CD138^+$ enriched cell fractions. (B) Tumor purity estimated from the exome copy number or mutation data based on the absolute allele frequency of constitutional variants in deletion regions or somatic SNV allele frequency in diploid regions of the genome when no usable deletions were detected in the tumor. (C) Distribution of observed somatic SNV allele frequencies.

### 4 Clinical and Molecular Associations with RNA Subtypes

#### 4.1 Mechanisms of RB1 Complete Loss at Diagnosis

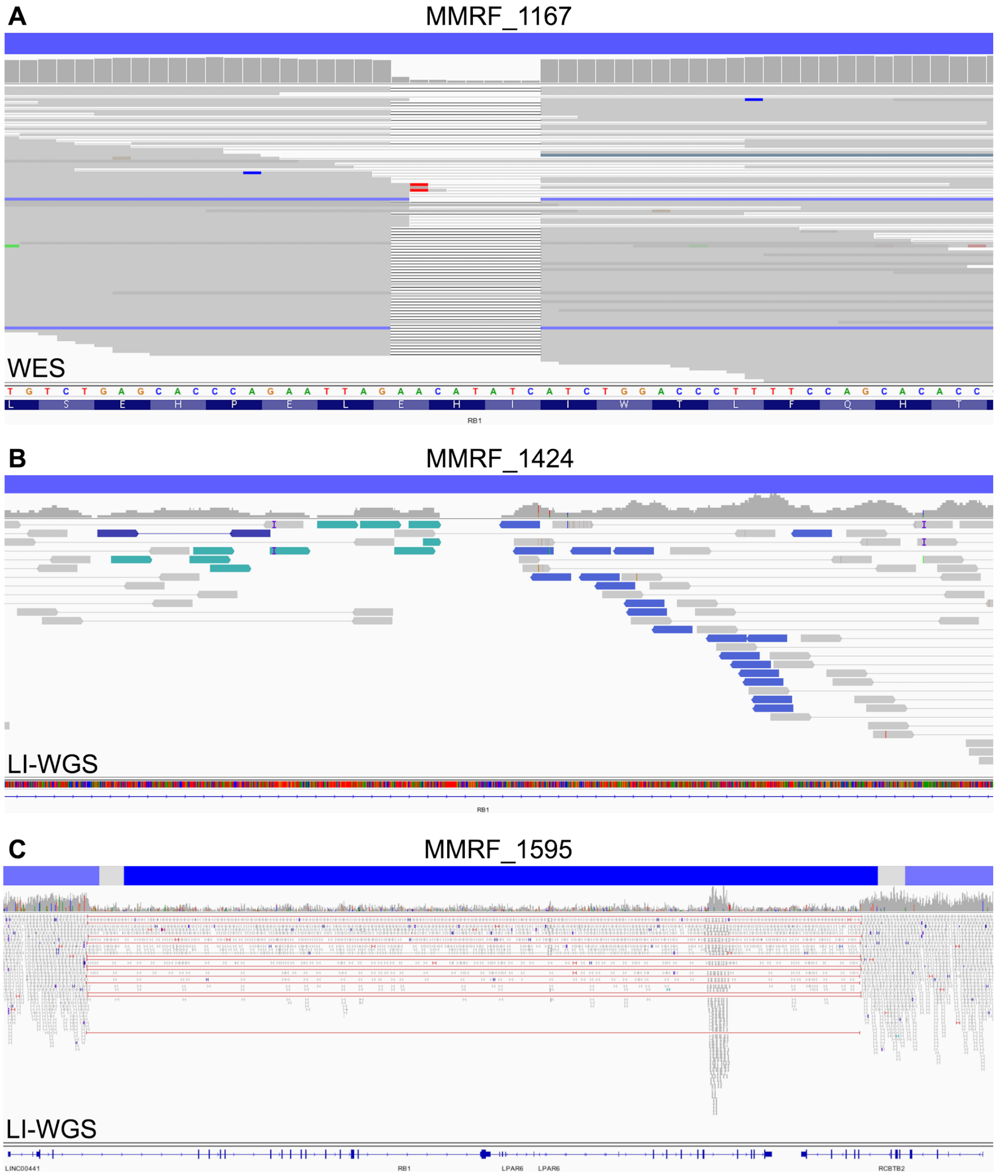


Different mechanisms of complete loss of RB1 are observed in tumors of the PR RNA subtype, but generally involve a one copy deletion of 13q coupled with a second molecular event. Panels A-C show CN segmentation (blue bars) and sequencing data (WES or LI-WGS) for three tumors of the PR RNA subtype at baseline. (A) Patient MMRF_1167 had complete loss of RB1 as a result of 13q copy loss (log2 CN = -1.004) and a small deletion (AR = 0.87). (B) Patient MMRF_1424 had complete loss of RB1 as a result of 13q copy loss (log2 CN = -0.9708) and an inversion where the breakpoint in the intronic region between exons 2 and 3 prevents splicing. (C) Patient MMRF_1595 had complete loss of RB1 as a result of 13q copy loss (log2 CN = -1.006) and a second interstitial deletion (log2 CN = -5.3759) including all RB1 exons except exon 1.

### 5 Transition to PR at Progression and Link with G1/S

#### 5.1 Change in RNA Subtype Probabilities Over Time

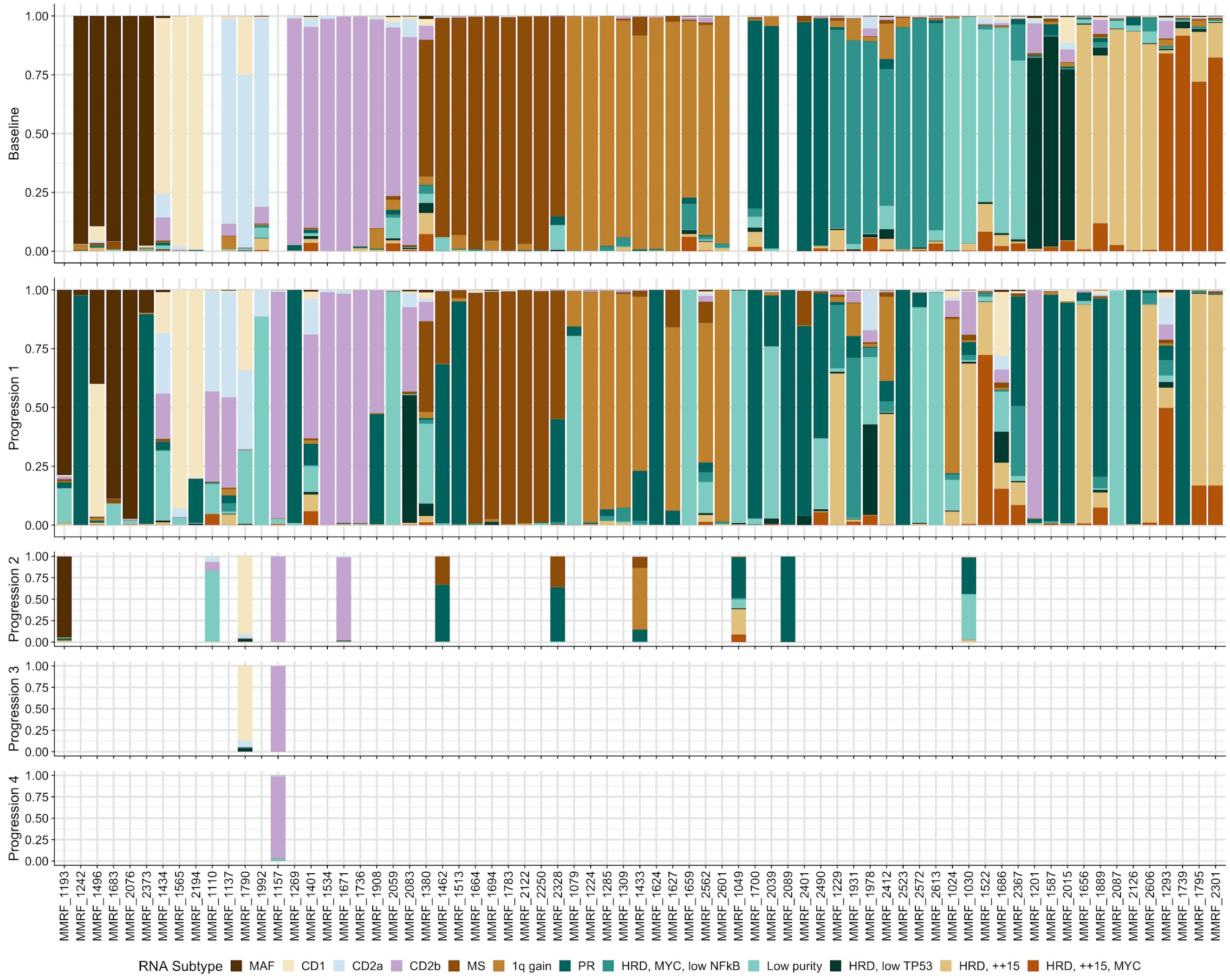


RNA subtype probabilities for the 71 serial patients with RNAseq data at two or more timepoints. All patients classified in the low purity subtype at baseline have a discernable RNA subtype other than low purity at progression, supporting the observation that this subtype is driven by sample purity. Shifts from a non-PR baseline subtype to a largely PR subtype or partial population of PR cells are evident.

#### 5.2 Overall Survival of Patients After Transition to PR Subtype

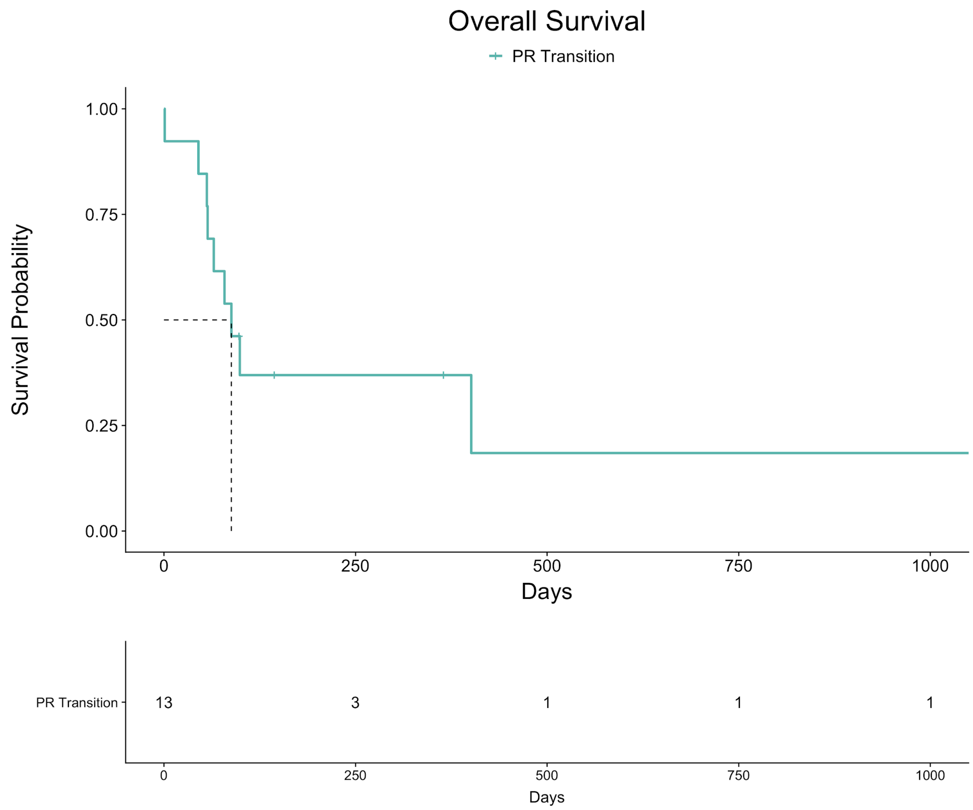


OS outcome for the 13 patients that transition to the PR subtype at progression from any non-PR RNA subtype at baseline, excluding the low purity subtype. Days are landmarked to the date at which the progression visit bone marrow sample was obtained to the date of last follow up (no OS censor flag, 4 patients) or to the date of death (OS censor flag, 9 patients). Patients that transitioned to the PR subtype exhibited extremely poor survival outcomes, with median OS of 88 days (3 months) after the progression visit.

#### 5.3 Deletion of of CDKN2C in Patients that Transitioned to PR

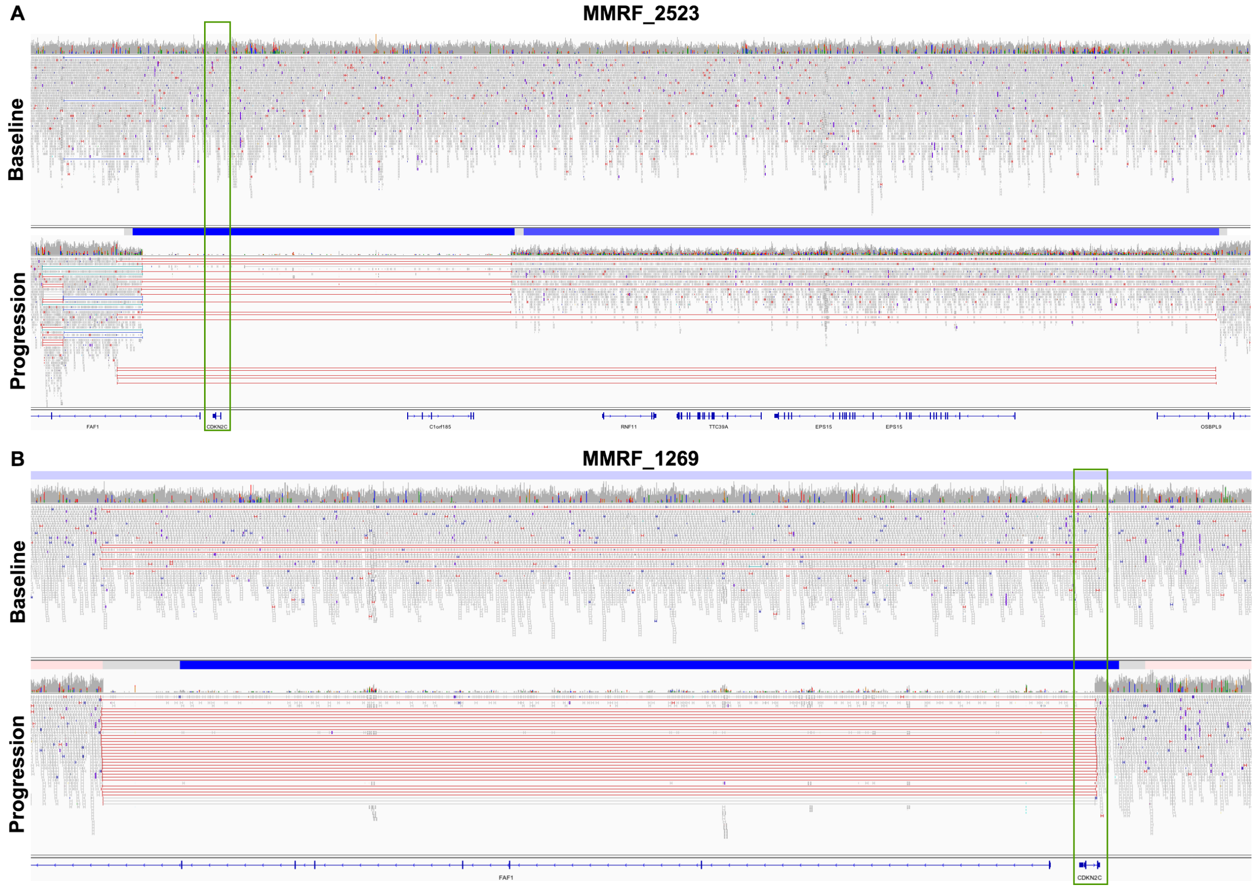


Two patients that transitioned to the PR subtype at progression acquired complete loss of function of CDKN2C due to overlapping deletion. Panels show long-insert WGS reads from tumor samples for patients MMRF_2523 (A) and MMRF_1269 (B) at baseline (non-PR) and progression (PR). (A) At baseline, patient MMRF_2523 was diploid (log2 CN = -0.0747) with no evidence of a deletion spanning CDKN2C however, at progression the patient had a 2-copy deletion of CDKN2C (blue bar, log2 CN = -3.3505) due to two unique deletions (red bars) spanning CDKN2C (green box). (B) At baseline, patient MMRF_2523 had a 1 copy loss of CDKN2C (light blue bar, log2 CN = -0.3511) due to a larger deletion on chr1. There is also read evidence supporting a deletion involving CDKN2C/FAF1, suggesting that a subclonal population with complete loss of CDKN2C was present at diagnosis in this patient. At progression, when the patient transitioned to PR, the patient's tumor had a 2-copy deletion of CDKN2C (dark blue bar, log2 CN = -4.6212). In this patient, the minor clone harboring the CDKN2C deletion at baseline constitutes the bulk of the tumor population at progression.

#### 5.4 Deletion of of CDKN1B in Patient that Transitioned to PR

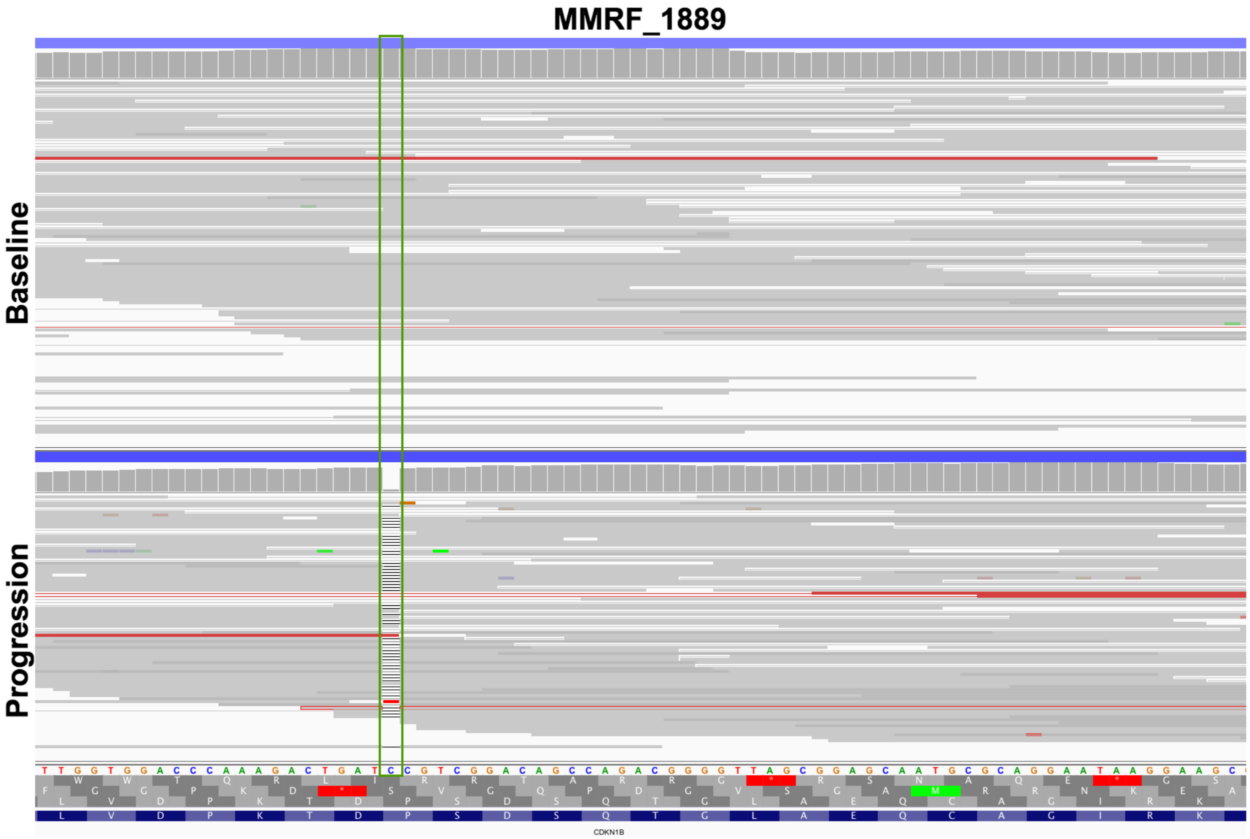


One patient that transitioned to the PR subtype at progression acquired complete loss of function of CDKN1B due to copy loss and mutation. Panels A and B show WES data for patient MMRF_1889 at (A) baseline and (B) progression, when the patient transitioned to the PR subtype. (A) At baseline the patient had a one copy loss of CDKN1B due to an arm-level deletion of 12p. (B) At progression, the patient had complete loss of CDKN1B due to copy loss of 12p and a clonal frameshift mutation (AR = 0.98). There was no read evidence supporting the existence of a subclone with this mutation at diagnosis, suggesting that this mutation was acquired.
